# Adverse Long-Term Outcomes and an Immune Suppressed Endotype in Sepsis Patients with Reduced Interferon-γ ELISpot: A Multicenter, Prospective Observational Study

**DOI:** 10.1101/2023.09.13.23295360

**Authors:** Evan A. Barrios, Monty B. Mazer, Patrick McGonagill, Christian B. Bergmann, Michael D. Goodman, Robert W. Gould, Mahil Rao, Valerie Polcz, Ruth Davis, Drew Del Toro, Marvin Dirain, Alexandra Dram, Lucas Hale, Mohammad Heidarian, Tamara A. Kucaba, Jennifer P. Lanz, Ashley McCray, Sandra Meszaros, Sydney Miles, Candace Nelson, Ivanna Rocha, Elvia E Silva, Ricardo Ungaro, Andrew Walton, Julie Xu, Leilani Zeumer-Spataro, Anne M. Drewry, Muxuan Liang, Letitia E. Bible, Tyler Loftus, Isaiah Turnbull, Philip A. Efron, Kenneth E. Remy, Scott Brakenridge, Vladimir P. Badovinac, Thomas S. Griffith, Lyle L. Moldawer, Richard S. Hotchkiss, Charles C. Caldwell

## Abstract

**Background:** Sepsis remains a major clinical challenge for which successful treatment requires greater precision in identifying patients at increased risk of adverse outcomes requiring different therapeutic approaches. Predicting clinical outcomes and immunological endotyping of septic patients has generally relied on using blood protein or mRNA biomarkers, or static cell phenotyping. Here, we sought to determine whether functional immune responsiveness would yield improved precision.

**Methods:** An *ex vivo* whole blood enzyme-linked immunosorbent (ELISpot) assay for cellular production of interferon-γ (IFN-γ) was evaluated in 107 septic and 68 non-septic patients from five academic health centers using blood samples collected on days 1, 4 and 7 following ICU admission.

**Results:** Compared with 46 healthy subjects, unstimulated and stimulated whole blood IFNγ expression were either increased or unchanged, respectively, in septic and nonseptic ICU patients. However, in septic patients who did not survive 180 days, stimulated whole blood IFNγ expression was significantly reduced on ICU days 1, 4 and 7 (all p<0.05), due to both significant reductions in total number of IFNγ−producing cells and amount of IFNγ produced per cell (all p<0.05). Importantly, IFNγ total expression on day 1 and 4 after admission could discriminate 180-day mortality better than absolute lymphocyte count (ALC), IL-6 and procalcitonin. Septic patients with low IFNγ expression were older and had lower ALC and higher sPD-L1 and IL-10 concentrations, consistent with an immune suppressed endotype.

**Conclusions:** A whole blood IFNγ ELISpot assay can both identify septic patients at increased risk of late mortality, and identify immune-suppressed, sepsis patients.

**Trial Registry:** Because the study is a prospective observational study, and not a clinical trial, registration with *clinical trials.gov* is not required.

## Introduction

Sepsis remains one of the most common causes of critical illness and too often leads to death and morbidity (1-3). Importantly, sepsis is a pathophysiologic host response to microbial infection associated with organ injury and dysfunction (4). However, the nature and magnitude of the host response to sepsis is highly variable depending upon the subject’s age, comorbidities and source and severity of microbial infection. Although sepsis is frequently associated with an early exaggerated inflammatory response (5), persistent inflammation (6,7), coagulopathy (8), prolonged immune suppression (3,6,9-12), and lean tissue wasting (13,14), the contribution of these individual responses to the overall outcome of the patient is still unresolved (15,16). Precision medicine has been proposed as a tool to identify which immunologic endotype drives organ injury and is an appropriate target for therapeutic intervention (17). Biomarkers, based on static blood cell phenotypes, protein and transcriptomic metrics have been commonly used to endotype critically ill patients with and without sepsis (15,17-20).

Enzyme-Linked ImmunoSpot assay (ELISpot) is a widely used immunological technique that enables the detection and quantification of individual cells responding to external receptor-specific and nonspecific stimulants and secreting specific proteins, particularly cytokines (21). This method is important for studying the immune response at the single-cell level, offering valuable insights into immune cell function, and immune-related diseases. Its ability to analyze immune responses at the cellular level makes it particularly suitable for monitoring immune system functionality in sepsis. In the current report, we examined the extent to which whole blood ELISpot production of interferon γ (IFNγ) can identify immune suppressed, critically ill patients at increased risk of death.

## Materials and Methods

This multi-center, prospective diagnostic and prognostic study conducted between February 23, 2021, and July 22, 2022, enrolled two cohorts of critically ill patients at the time of ICU admission. The first cohort included patients with a suspected diagnosis of sepsis admitted to the ICU (SEPSIS). The second cohort included critically ill patients admitted to the ICU without currently suspected sepsis (Critically Ill, Non-Septic; CINS), but considered at high risk for subsequent infection (e.g., postoperative, severe trauma). Patient enrollment is shown in **Figure 1**, consistent with Enhancing the Quality and Transparency of Health Research Standards for Reporting of Diagnostic Accuracy (STARD) reporting guidelines (22). All patients were managed under institutional clinical management protocols. Centralized ethics approval was obtained from the University of Florida Institutional Review Board (#IRB 202000924) which served as the sponsoring institution. Written informed consent was obtained from each patient or their proxy decision-maker at individual clinical sites.

**Figure 1.**
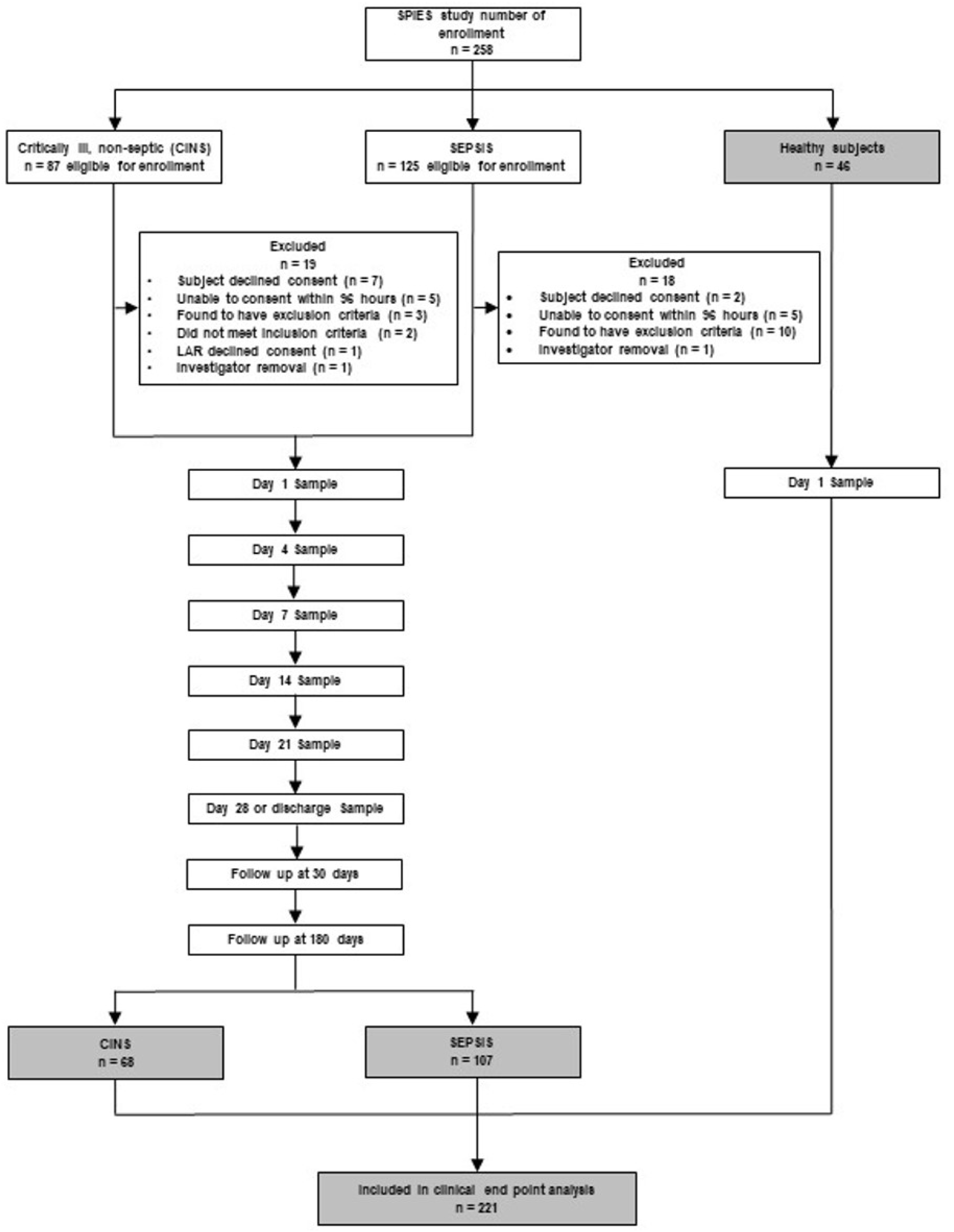
Flow Diagram for Study Enrollment.

A heparinized blood collection tube (Becton Dickinson) was obtained within the first three days of ICU admission (labeled as day 1), and on subsequent days 3 through 5 (labeled as day 4), and weekly thereafter (± 2 days). Self-or proxy-reported race and ethnicity category data were collected as per National Institutes of Health reporting guidelines and requirements.

Inclusion criteria consisted of ICU admission from the emergency department for community-acquired sepsis or severe trauma (patients with Injury Severity Scores >15, hemorrhagic shock, and/or severe chest trauma), as well as non-trauma, postoperative ICU admission, ICU transfer from the emergency department and inpatient transfer from ward to ICU.

Sepsis was defined according to Sepsis-3 criteria (4), and all subjects were clinically adjudicated at the individual participating sites. Patients admitted to the ICU for critical illness, non-sepsis (CINS) were also adjudicated to rule out sepsis. A detailed summary of inclusion and exclusion criteria is provided in the **Supplemental Materials and Methods.**

Healthy control subjects were recruited at each of the clinical sites. Efforts were made to age, sex and race-ethnicity match the healthy control subjects to the SEPSIS cohort. Individuals with autoimmune diseases being treated with biologic immune modulators were excluded, as were individuals who had received anti-neoplastic therapies or diagnosed with cancer within the past six months. Vulnerable populations were also excluded.

### Primary Outcomes and Clinical Adjudication

The primary clinical outcome for ELISpot was 180-day mortality, determined via clinical records and telephone follow-up with the patient, their proxy, or their designated contact, and cross-checked through the US Social Security Death Index. We analyzed temporal trends of ELISpot in both SEPSIS and CINS patients but compared estimated performance of predictive models primarily in the SEPSIS patients, as 180-day mortality in the CINS cohort was <4%. Final sepsis or CINS adjudication was performed by physician-investigator determination at each clinical site at completion of each patient’s hospital course.

Secondary clinical outcome variables included all-cause (in hospital, 30-day) mortality, development of chronic critical illness (CCI), secondary infections, and poor discharge disposition. Inpatient clinical trajectory was defined as “early death”, “rapid recovery”, or “CCI”. CCI was defined as an ICU length of stay ≥14 days with evidence of persistent organ dysfunction (SOFA score ≥ 2) (23). Hospitalized patients who died after an ICU length of stay >14 days from the index hospitalization were also classified as CCI. Poor disposition was defined as discharge to a skilled nursing facility (SNF), long-term acute care facility (LTAC), or hospice. Secondary infections were defined as per the US Centers for Disease Control and Prevention criteria.

### ELISpot

ELISpot assays were conducted using the human IFNγ Immunospot^®^ kit (CTL Inc., Cleveland, OH) with several important modifications including the use of diluted whole blood as previously described (21). Specifically, 100 μl of heparinized whole blood was diluted 1:10 with kit buffer and 50 μl of the diluted sample was added to each well. Samples were incubated in wells containing either buffer alone or a soluble anti-CD3/anti-CD28 (125 ng/ml/1.25 μg/ml) mAb agonist (BioLegend, San Diego, CA). Samples were assayed in duplicate. Optimal concentrations of agonist were determined in preliminary studies (see **Supplemental Materials and Methods**).

Samples were quantitated using a CTL S6 Entry or S6 FluoroCore™ ELISpot reader at each clinical site. To assure comparable results, the instruments were harmonized by CTL Inc. prior to study using an external standard across all five clinical sites. Results are presented as the number of spot-forming units (SFU), spot size (μm^2^; SS) and total expression (μm^2^; TE) a product of the number of spots and mean spot size using the Immunospot^®^ SC software suite (version 7.0.30.4). Spot forming units represent individual blood cells expressing IFNγ and spot size is an indication of the amount of IFNγ produced per cell. In subsequent analyses, the number of IFNγ-producing cells was adjusted for each individual patient’s absolute lymphocyte count to yield the percentage of total lymphocytes expressing IFNγ.

### Additional Laboratory Analyses

Whole blood total leukocyte and absolute lymphocyte counts were determined on EDTA-anticoagulated whole blood at the individual clinical sites either using their hospital’s Clinical and Diagnostics Laboratory or a research Beckman-Coulter Dx500 or Dx900 hemocytometer (Brea, CA). Cytokine and additional plasma protein analyses were conducted at the University of Florida Sepsis and Critical Illness Research Center (SCIRC) where they were determined in batch using the Luminex MagPix^®^ (Austin, TX) platform using commercial reagents.

Excess sample was stored at the Biorepository of the Clinical and Translational Science Institute (https://www.ctsi.ufl.edu/research/laboratory-services/ctsi-biorepository-2/) where it is available to the scientific community under guidelines promulgated by the National Institute of General Medical Sciences.

### Data Collection and Analysis

Clinical data collection was conducted at each site and entered into a web-based electronic case report form created on the REDCap™ platform managed by the University of Florida Clinical and Translational Science Institute (CTSI). Access to the case report form was password protected and limited to only approved research staff, and all interactions with the database were recorded. Peer-to-peer communication allowed approved individuals at all five sites access to their own data and de-identified data from the other four clinical sites. Research data, including ELISpot, total leukocyte and absolute lymphocyte counts, and plasma protein and cytokine data were uploaded into the case report forms from the University of Florida SCIRC. Data managers at the SCIRC were responsible for creating final locked datasets for subsequent analysis.

### Statistical Analysis

Descriptive data are presented as frequencies and percentages or medians and IQRs. The Fisher exact test and Mann-Whitney or Kruskal-Wallis ANOVA tests were used for categorical and continuous variables, respectively. Area under the receiver operating characteristics curve (AUROC) values with 95% confidence intervals (computed with 2000 stratified bootstrap replicates) were used to assess discrimination. Univariable and multivariable logistic regression were performed to assess whether the combination of metrics improved overall performance. Post hoc tests were performed for continuous outcomes using the Dunn test. For post hoc analyses of categorical outcomes, separate 2 × 2 Fisher exact tests were performed. All significance tests were 2-sided, with a raw p ≤ 0.05 considered statistically significant.

Analyses were performed using the R Project statistical package, version 4.1.0 (R Project for Statistical Computing).

## Results

### Patient Characteristics

Demographic characteristics of the 175 enrolled patients (99 men [57%] and 76 women [43%]) and 46 healthy control subjects (16 men [36%], 30 women [64%]) are summarized in **Table 1**. The overall cohort included 107 and 68 patients in the SEPSIS and CINS cohorts, respectively. Patient characteristics were similar across all three cohorts with the exception that the healthy control subjects were more predominantly female and younger, and SEPSIS patients had a higher Charlson comorbidity index than CINS subjects (**Table 1**). Within the CINS cohort ([n□=□68]), reason for ICU admission was identified in **Supplemental Table 1**.

**Table 1.**
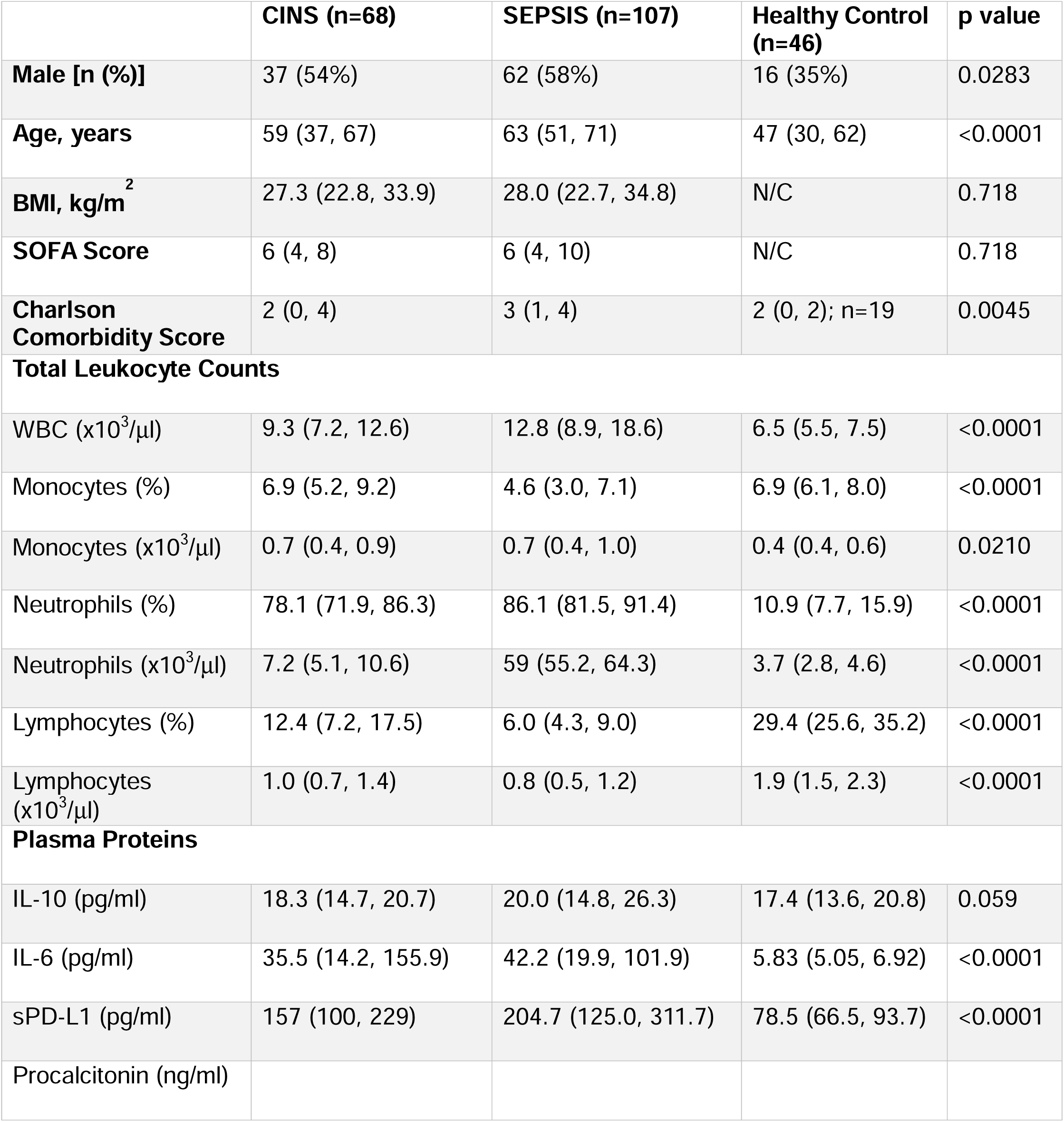
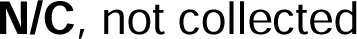
Clinical Characteristics of Patient Cohorts and Healthy Control Subjects. Patients included two cohorts of individuals admitted to the ICU, one presumed to be septic (SEPSIS), a second presumed to not be septic, but critically ill and at risk of developing sepsis (CINS). Values are obtained at time of admission to the ICU.

**Table 2** shows clinical outcomes for the SEPSIS and CINS patients. Hospital length of stay (p<0.02), incidence of secondary infections (p<0.001), development of CCI (p<0.001), and in-hospital mortality (p<0.01) were all significantly higher in SEPSIS than in CINS patients.

**Table 2.**
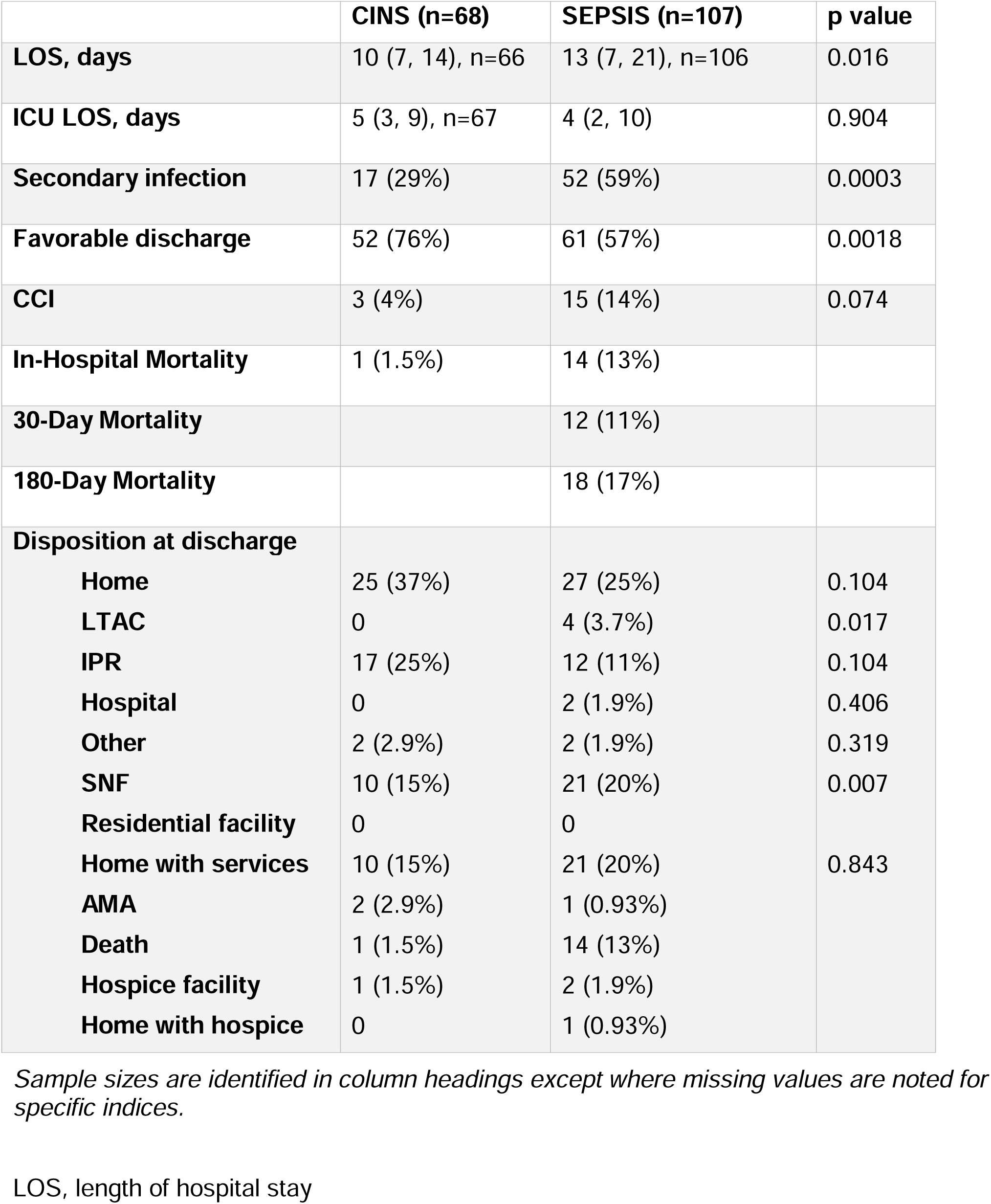
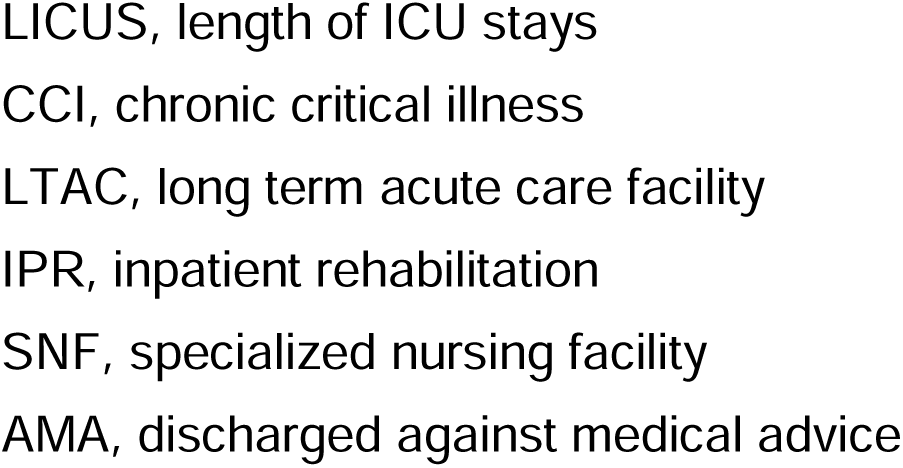
Clinical Course and Clinical Outcomes in SEPSIS and CINS cohorts.

Disposition at discharge also significantly differed between SEPSIS and CINS patients, as did 30- and 180-day mortality (both p<0.01).

### IFN**γ** Production by Unstimulated and anti-CD3/CD28 mAb-Stimulated Whole Blood

Comparison of the first sample collected (days 1-3 post ICU admission) among the three cohorts revealed considerable heterogeneity in the individual subject response, irrespective of the cohort. Surprisingly, as a group, spontaneous IFNγ production in *unstimulated* whole blood was significantly increased from SEPSIS and CINS patients than from healthy subjects at all sampling intervals (days 1, 4 and 7), despite a significant reduction in lymphocyte numbers (**Figure 2, Supplemental Figure 1**). This was reflected generally by an increased number of IFNγ-producing cells (SFU) (all p<0.05;), although the amount of IFNγ produced by each cell (SS) was increased on day 4 (**Figure 3**). When the total number of IFNγ producing cells was adjusted for the absolute lymphocyte count, the percentage of lymphocytes expressing IFNγ was further increased significantly in both SEPSIS and CINS (p<0.001; **supplemental Figure 2**). There was no difference between unstimulated IFNγ expression between the SEPSIS and CINS cohorts.

**Figure 2.**
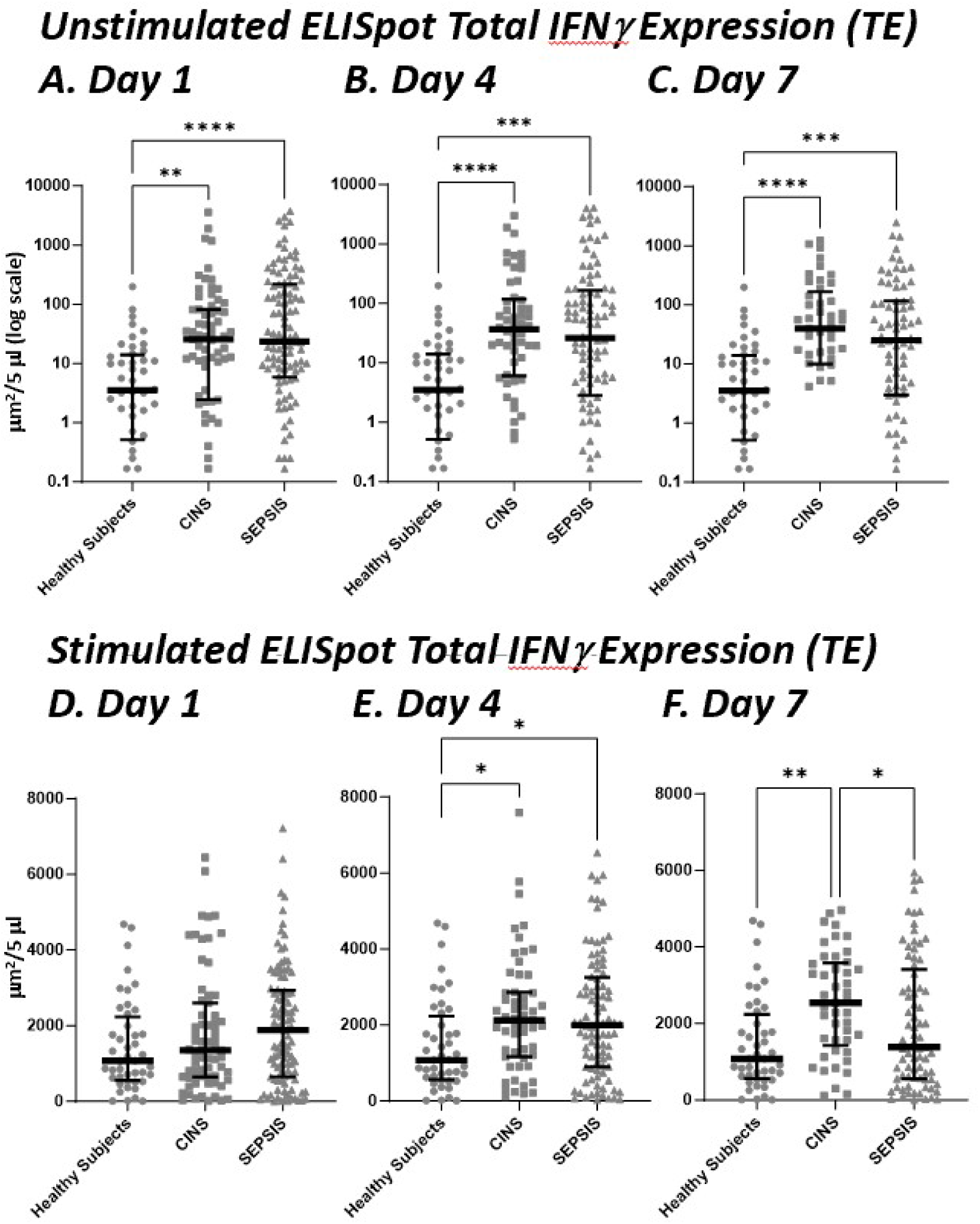
Unstimulated and Stimulated IFNγ Expression as Determined by ELISpot in SEPSIS and CINS Patients, and Healthy Control Subjects on Days 1, 4, and 7 Following ICU Admission. Values represent medians and individual subject responses. Panels A, B, C are from unstimulated whole blood while Panels D, E, F are antiCd3/CD28 stimulated whole blood. Note that the scales for unstimulated expression are logarithmic, whereas they are linear for stimulated expression to appropriately reflect the magnitude and heterogeneity of the individual response. * p< 0.05, ** p<0.01, ***p<0.001, ****p<0.0001 as determined by Kruskal-Wallace ANOVA and post-hoc analyses using the Dunn test. Values are two sided and represent raw p values. SFU, spot forming units. SS, spot size. TE, total IFNγ expression.

**Figure 3.**
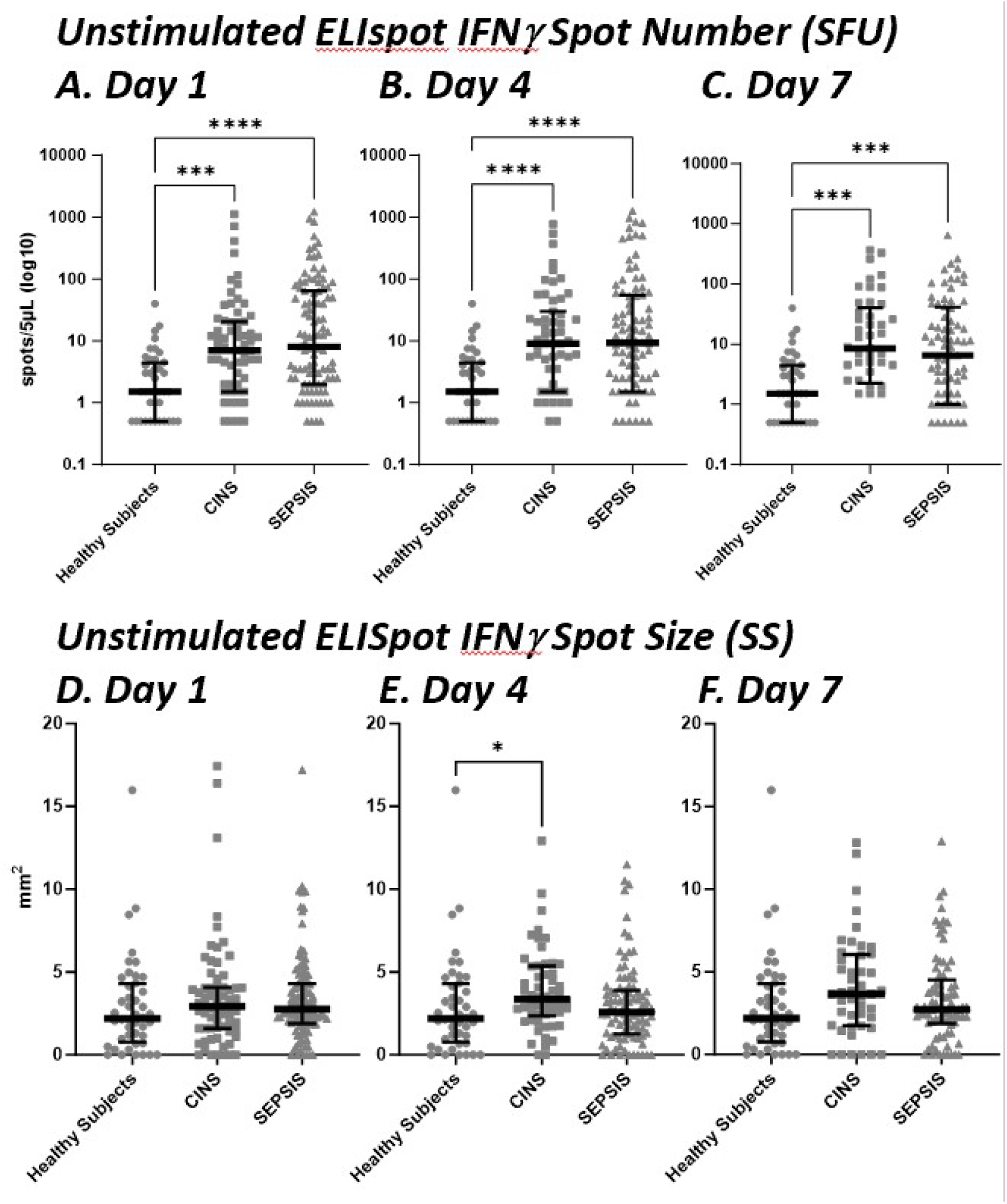
– ELISpot Spot Forming Units (SFU) (Panels A, B, C) and Spot Size (SS) (Panels D, E, F) from Unstimulated Whole Blood in the Three Cohorts (Healthy Subjects, SEPSIS and CINS). In unstimulated whole blood, SEPSIS and CINS cohorts demonstrated a consistent increase in the number of cells (SFU) producing IFNγ, when compared to healthy subjects. * p<0.05, *** p<0.001, **** p<0.0001, as determined by Kruskal-Wallace ANOVA and post-hoc analyses using the Dunn test. Values are two sided and represent raw p values. SFU, spot forming units. SS, spot size. TE, total IFNγ expression.

*Ex vivo* stimulation of the whole blood from the three cohorts with agonist anti-CD3/CD28 mAb resulted in expected increases in the total expression of IFNγ produced when compared with unstimulated samples. There were also increase in the total amount of IFNγ produced per unit volume of blood on days 4 and 7 when comparing CINS to healthy subjects (**Figure 2**). This increased IFNγ expression was only seen on day 4 in SEPSIS patients.

### Influence of Outcome on ELISpot Responses

SEPSIS subjects had a greater in-hospital, 30- and 180-day mortality when compared to CINS patients (**Table 2).** In addition, the incidence of secondary infections, development of CCI and an adverse discharge disposition were all significantly greater in SEPSIS than CINS patients (all p<0.05).

SEPSIS subjects who died within 180 days of ICU admission did not differ from surviving patients based on their admission or day 1 SOFA scores or total leukocyte numbers, even though non-surviving SEPSIS patients were significantly older and had higher Charlson comorbiditiy scores (both p<0.05; **Supplemental Table 1**). Interestingly, there were marked differences in the IFNγ production from stimulated whole blood between sepsis subjects who survived or did not survive 180 days. Both the absolute number (SFU) and percentage of IFNγ producing cells, and the spot size (SS) were significantly lower, and therefore, total expression (TE) was reduced in non-surviving versus surviving SEPSIS patients (all p<0.05; **Figure 4**).

**Figure 4.**
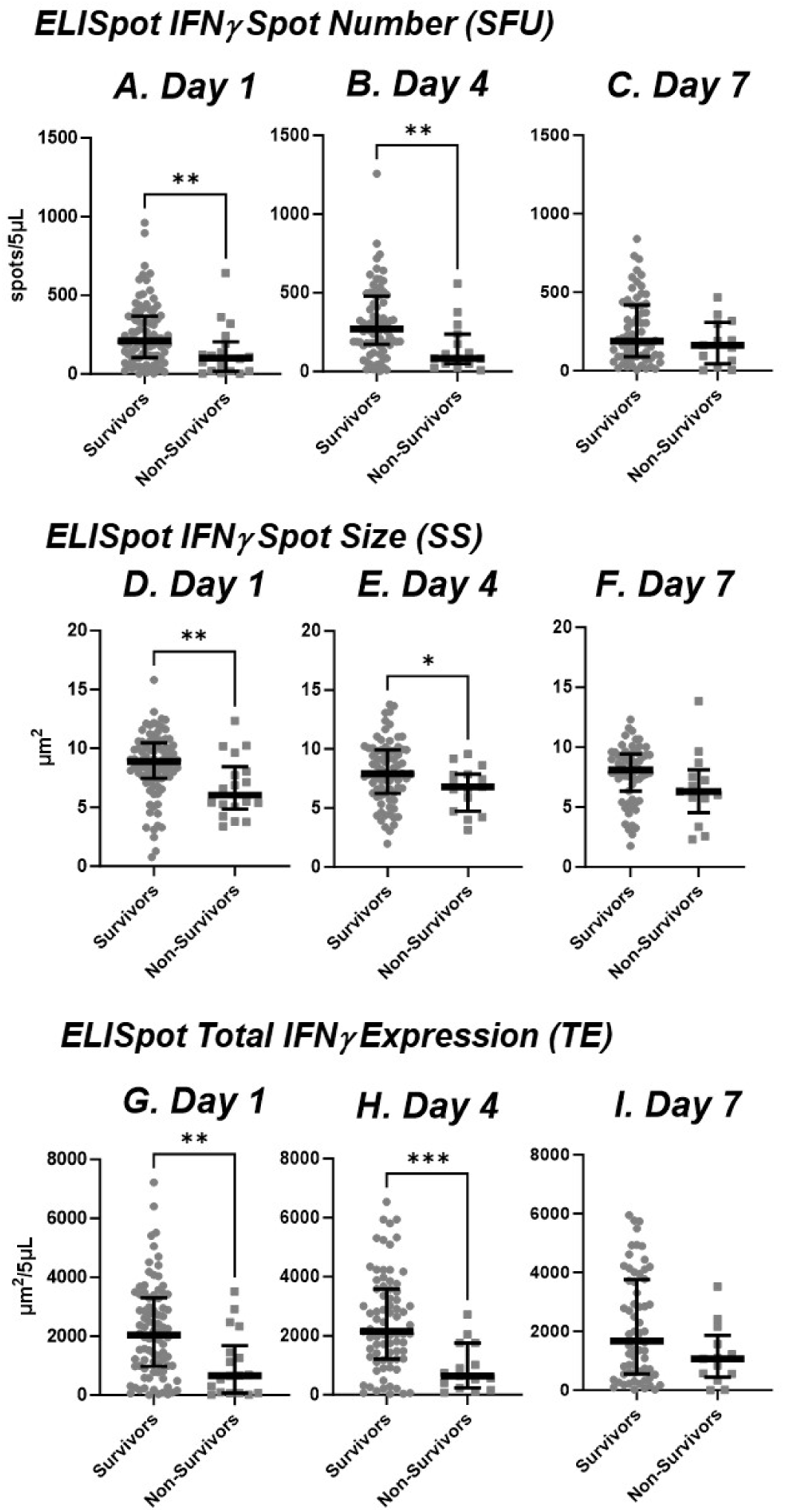
Anti-CD3/CD28 Stimulated IFNγ Expression by ELISpot in Sepsis Patients Measured at 1, 4 and 7 Days after ICU Admission who Survived or Did Not Survive by 180 days. Panels A, B, C reflect spot number, D, E, F, spot size, and G, H, I, total IFNγ expression. Values represent medians and individual subject responses. The number of subjects declines over time as patients are either discharged from ICU or die. * p< 0.05, ** p<0.01, ***p<0.001, ****p<0.0001 as determined by Kruskal-Wallace ANOVA and post-hoc analyses using the Dunn test. Values are two sided and represent raw p values. SFU, spot forming units. SS, spot size. TE, total IFNγ expression.

This reduction in expression was, in general, sustained through day 4 in non-survivors (**Figure 4**). After 7 days, there were too few ICU-remaining patients to continue the comparison (*data not shown*). Surprisingly, IFNγ production from *unstimulated* whole blood did not differ between surviving and non-surviving patients at any time point (*data not shown*). There also did not appear to be any significant changes in ELISpot responses (both stimulated and unstimulated) over time in either surviving or non-surviving individual SEPSIS patients (*data not shown*).

### Univariate and Multivariate Prediction Models for Long-term Survival and Secondary Outcomes

Because of the differences in IFNγ expression between surviving and non-surviving SEPSIS patients, ELISpot AUROCS were evaluated for their discriminatory prediction of long-term survival (180 day), as well as secondary outcomes, and compared to clinical indices (SOFA, Charlson comorbidity scores), total white blood cell and absolute lymphocyte counts, and plasma protein markers (selected cytokines, procalcitonin, and sPD-L1) in the SEPSIS patients. Similar combined SEPSIS and CINS analyses could not be performed due to the low mortality in CINS patients. Results are presented in **Table 3** and **Figure 5**.

**Figure 5.**
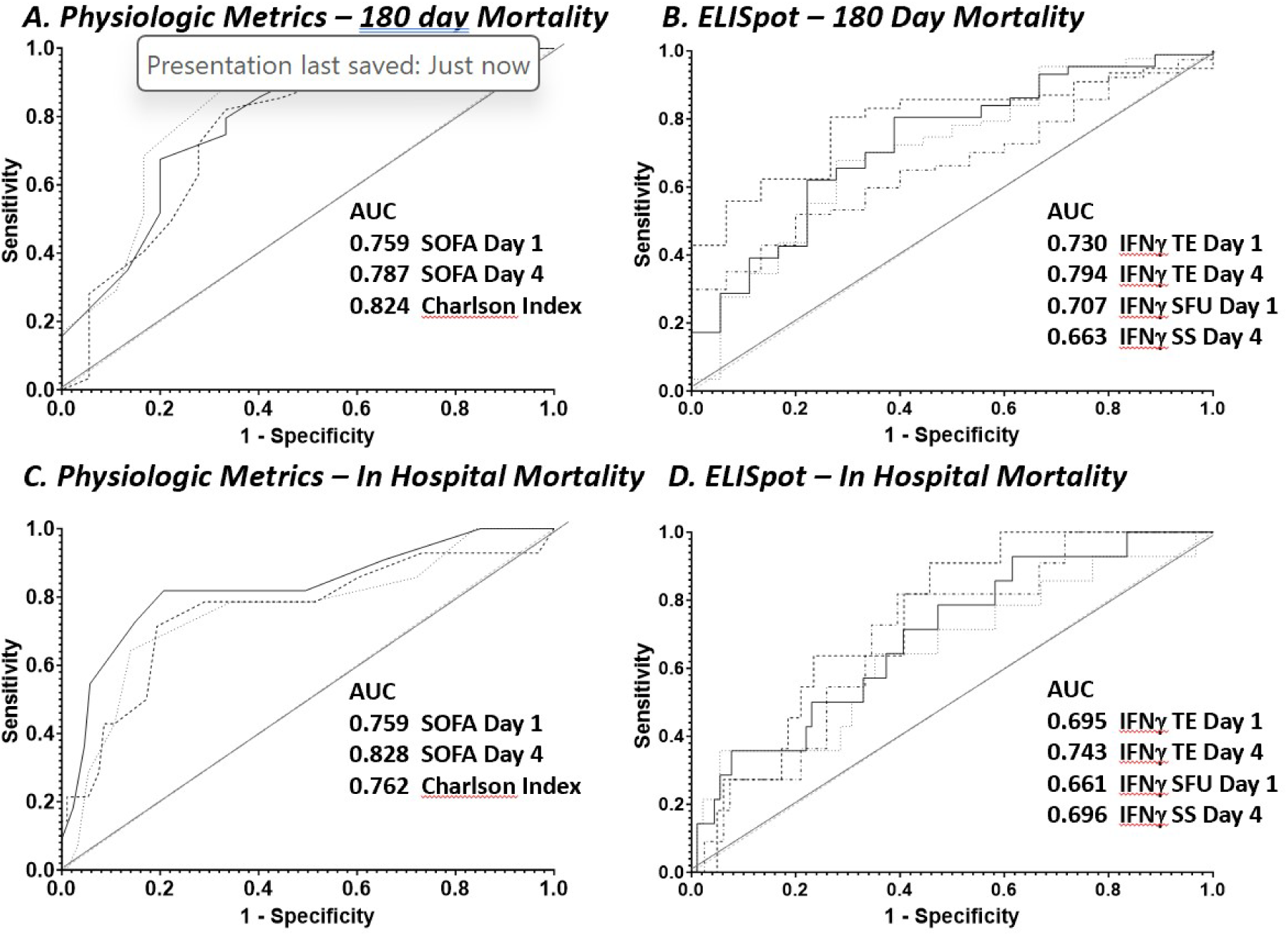
Area under the Receiver Operator Curves (AUROC) for Physiologic (SOFA, Charlson comorbidity scores) and Stimulated IFNγ ELISpot Responses in Differentiating In-Hospital and 180-day Mortality. Top Row: Panel A provides SOFA and Charlson Comorbidity Index, and Panel B, selected ELISpot parameters discriminating 180 day mortality. Lower Row: Same breakdowns as for the top row discriminating in-hospital mortality. TE, IFNγ ELISpot total expression; SFU, IFNγ ELISpot spot forming units; SS IFNγ ELISpot spot size

**Table 3.**
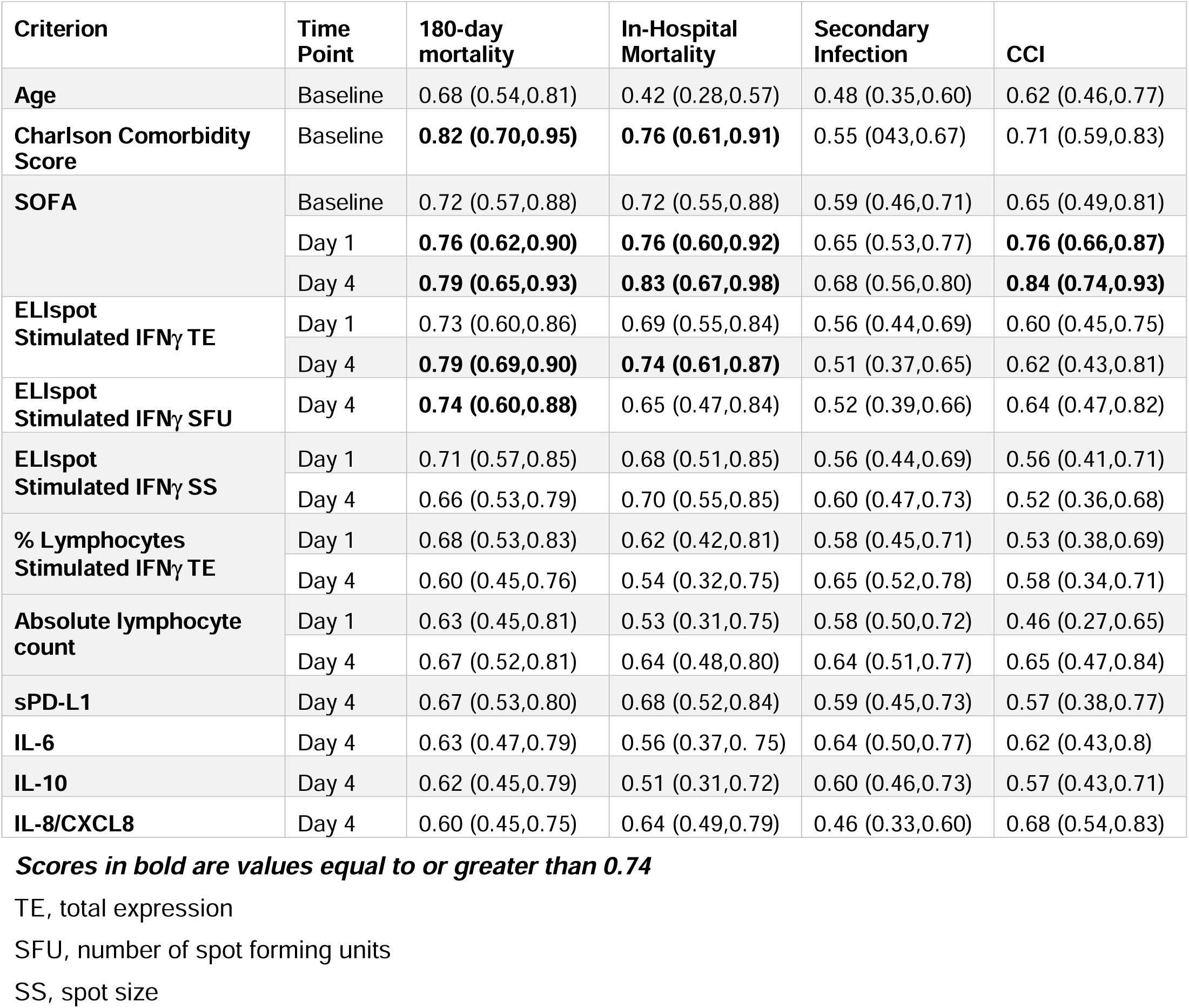
Selected Area Under the Receiver Operating Curve Discrimination for Primary Outcome Variables in the SEPSIS cohorts. Values represent the mean and 95% confidence intervals for the primary outcome variables: 180-Day Mortality, In-Hospital Mortality, Incidence of Secondary Infection, Development of Chronic Critical Illness (CCI).

Similar to data reported by ourselves and others (24-26), the most consistent discriminator of 180-day mortality was the Charlson comorbidity score (AUROC; 0.824 [CI 0.700-0.948], which also discriminated in-hospital mortality (0.762 [0.611-0.912]) and development of CCI (0.713 [0.594-0.832]), but not the incidence of secondary infection (0.549 [0.426-0.673]. Importantly, stimulated total IFNγ expression at day 1 and day 4 did not significantly differ from the Charlson comorbidity score with AUROCs of 0.730 (0.601-0.859) and 0.794 (0.691-0.897), respectively. Total IFNγ expression on day 4 also discriminated in-hospital mortality with an AUROC of 0.743 (0.615-0.830), but it was not a strong discriminator of either development of CCI or incidence of secondary infections.

For total stimulated IFNγ expression, the components contributing to its discriminative power for 180-day mortality were both the number of IFNγ producing cells (SFU; Day 1, 0.707 [0.568- 0.846] and the amount of IFNγ produced by individual cells (SS; Day 1, 0.707 [0.568-0.846]).

The discriminatory power of the ELISpot total expression (TE) for predicting both in-hospital and 180-day mortality was much greater than seen for either absolute lymphocyte count (ALC), IL-6, or procalcitonin at either day 1 or day 4, or for changes in these parameters between days 1 and 7 (**Table 3**). In addition, the last ELISpot measurement obtained from the patient prior to discharge or death (usually day 7 or later) was also found to be not as discriminatory as the earlier day 1 and 4 measurements (*data not shown*).

Setting the threshold for day 1 and day 4 stimulated total IFNγ expression at approximately 80% sensitivity to discriminate 180-day survival, it was possible to assess the immunosuppressive endotype of those SEPSIS patients with reduced ELISpot total expression (**Table 4**). These individuals were older and had absolute lymphocyte counts significantly lower at both days 1 and 4 than SEPSIS patients above the ELISpot threshold. In addition, plasma sPD-L1 concentrations were significantly higher on day 4. Patients below the ELISpot threshold also had in-hospital and 180-day mortality, and development of CCI at a markedly higher frequency than those individuals above the threshold (all p<0.05).

**Table 4.** Evidence of Immunosuppression in SEPSIS Patients with Reduced Stimulated IFNγ Total Expression (TE) As Discriminated by ELISpot at Days 1 and 4 Post-ICU Admission. The threshold for reduced TE was set at 80% sensitivity. On day 1 the specificity was 42.1% and on day 4 the specificity was 63.2%.

| **Day 1** | **ELISpot IFNγ TE**<br>**<2334/$\mu\text{m}^2$**<br>**N=65** | **ELISpot IFNγ TE ≥2334 ($\mu\text{m}^2$)**<br>**N=40** | **P-value** |
| --- | --- | --- | --- |
| Age | 64.2 (14.7) | 54.6 (15.1) | 0.002 |
| Total WBC cell<br>day 1 ($\times 10^3$ /ml) | 13.9 (7.07) | 15.7 (7.77) | 0.158 |
| Absolute<br>Lymphocyte<br>Count Day 1<br>(/$\mu\text{l}^3$) | 980 (249) | 1460 (600) | 0.028 |
| sPD-L1 day 1<br>(pg/ml) | 315 (333) | 263 (246) | 0.311 |
| IL-10, day 1<br>(pg/ml) | 32.3 (70.0) | 28.6 (37.7) | 0.608 |
| **Outcome** |  |  | **Odds-ratio** |
| 180-day<br>mortality | 23.1% | 7.5% | 3.70<br>(1.00,13.7) |
| In-Hospital<br>mortality | 18.5% | 5.0% | 4.30<br>(0.91,20.3) |
| CCI | 18.5% | 7.5% | 2.79<br>(0.74,10.6) |
| Second<br>Infection | 59.6% (6<br>missing) | 58.8% (13 missing) | 1.03<br>(0.43,2.49) |

| **Day 4** | **ELISpot IFNγ TE**<br>**<1755/$\mu\text{m}^2$ N=41** | **ELISpot IFNγ TE**<br>**≥1755 ($\mu\text{m}^2$) N=51** | **P-value** |
| --- | --- | --- | --- |
| Age | 66.2(14.0) | 58.1(15.7) | 0.011 |
| Total WBC cell<br>day 1 ($\times 10^3/\text{ml}$) | **13.6(6.93)** | **15.4(8.18)** | 0.305 |
| Absolute<br>Lymphocyte<br>Count Day 1<br>($\text{ml}^3$) | 70.7 (197) | 191 (580) | **0.006** |
| sPD-L1 day 1<br>(pg/ml) | 337(248) | 239(290) | **0.007** |
| IL-10, day 1<br>(pg/ml) | 42.7(89.9) | 24.0(27.7) | 0.086 |
| **Outcome** |  |  | **Odds-ratio** |
| 180-day<br>mortality | 29.3% | 5.9% | 6.62<br>(1.72,25.5) |
| In-Hospital<br>mortality | 19.5% | 5.9% | 3.88<br>(0.96,15.7) |
| CCI | 22.0% | 7.8% | 3.30<br>(0.94,11.7) |
| Second Infection | 58.8% (12 missing) | 56.4% (7 missing) | 1.10<br>(0.44,2.80) |

Finally, to examine whether ELISpot could improve the discriminatory power of standard clinical indices (SOFA and Charlson comorbidity scores), single and multivariate logistic regression analyses were performed and the AUROCs of the models were calculated by a four-fold cross-validation procedure (**Table 5**). Model I was built on baseline Charlson comorbidity data and day 1 SOFA scores yielding AUROCs for 180-day mortality (0.911 [0.858-0.965]); the odds ratios show baseline Charlson comorbidity data and day 1 SOFA scores are significant predictors for 180-day mortality (**Table 5**). Model II was built on stimulated total IFNγ expression from Day 4 yielding AUROCs for 180-day mortality (0.794 [0.732-0.855]); the odds ratios show stimulated total IFNγ expression is a significant predictor for 180-day mortality (**Table 5**). Although ELISpot data show significant odds ratios in Model II, the addition of stimulated total IFNγ expression to the model built on standard score indices (Model III) did not significantly increase either the AUROC or the odds ratio for 180-day (0.915 [0.851-0.979] or in-hospital mortality (0.883 [0.801-0.964]), or development of CCI (0.861 [0.830-0.892]) (**Table 5**).

**Table 5.**
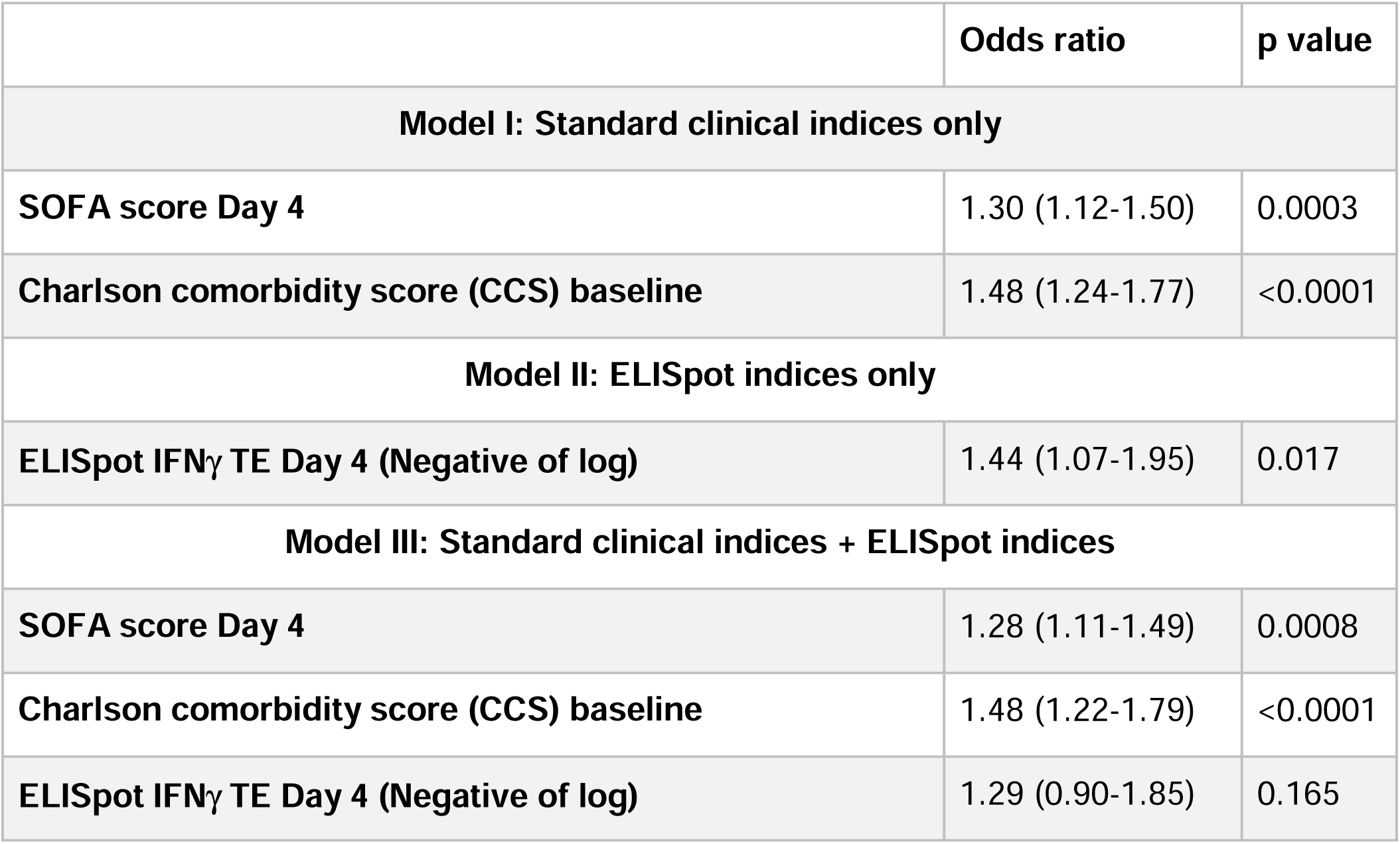
Multivariate Logistic Regression to Discriminate Time to Mortality based on Charlson Comorbidity and SOFA Scores and Day 4 ELISpot Stimulated IFNγ Total Expression in SEPSIS Patients.

|  | Odds ratio | p value |
| --- | --- | --- |
| **Model I: Standard clinical indices only** |  |  |
| **SOFA score Day 4** | 1.30 (1.12-1.50) | 0.0003 |
| **Charlson comorbidity score (CCS) baseline** | 1.48 (1.24-1.77) | <0.0001 |
| **Model II: ELISpot indices only** |  |  |
| **ELISpot IFNγ TE Day 4 (Negative of log)** | 1.44 (1.07-1.95) | 0.017 |
| **Model III: Standard clinical indices + ELISpot indices** |  |  |
| **SOFA score Day 4** | 1.28 (1.11-1.49) | 0.0008 |
| **Charlson comorbidity score (CCS) baseline** | 1.48 (1.22-1.79) | <0.0001 |
| **ELISpot IFNγ TE Day 4 (Negative of log)** | 1.29 (0.90-1.85) | 0.165 |

## Discussion

### Key Findings

This prospective, multi-center observational study has demonstrated that the adaptive immune response to critical illness, as defined by *ex vivo* whole blood production of IFNγ in response to T cell receptor stimulation, varied in response to critical illness (**Figures 2, 3**) and could also discriminate long-term outcomes (**Figure 5**, **Table 3**). Spontaneous IFNγ production by diluted whole blood was significantly increased in critically ill patients, irrespective of whether the critically ill patients were septic. In addition, when whole blood was stimulated *ex vivo* with a T cell receptor agonist, IFNγ production increased dramatically in both healthy and critically ill cohorts (both SEPSIS and CINS). However, stimulated ELISpot IFNγ total expression early in the admission to the ICU significantly differed between septic patients who survived 180 days and those who died, and this reduction in total expression in non-surviving septic patients was due to both reductions in the total number of IFNγ producing blood cells and the amount of IFNγ produced by individual cells (**Figure 4**). Importantly, day 1 and day 4 measurements were more discriminatory than later measurements. Using univariate modeling, stimulated ELISpot total expression measured in the first week of admission (sampling days 1 and 4) could differentiate 180-day mortality as well as SOFA and Charlson comorbidity scores, and markedly better than blood ALC, procalcitonin, IL-6 and sPD-L1 concentrations (**Table 3**). Septic patients who had low stimulated ELISpot total IFNγ expression at days 1 and 4 had an immunosuppressed endotype, as reflected in being older, and having lower ALC counts, higher plasma sPD-L1, and increased incidence of chronic critical illness, in-hospital and late mortality (**Table 4**). However, in multivariate models, stimulated ELISpot total IFNγ expression did not significantly improve the discrimination between 180-day survival in models built with SOFA and Charlson comorbidity scores (**Table 5**).

### Context

ELISpot has emerged as a powerful method to assess immunological status in a variety of clinical disorders (27-30), including sepsis and critically ill patients (20,21,31,32). It offers several theoretical advantages over other current metrics – for example, cell phenotypes (such as ALC (33-36) and HLA-DR expression on CD14^+^ blood cells (37,38)), plasma protein concentrations (such as procalcitonin (39-42), IL-6 (43-45) and sPD-L1 (46,47)) or blood transcriptomics (26,48,49) – used to predict both the severity of the host response and the immunosuppressed endotype. ELISpot, unlike these static measures, assesses one component of the functional status of the host protective immune response. In the present study, ELISpot revealed the capacity of T cells in the blood to produce IFNγ with and without stimulation through the T cell receptor.

This study is not the first to demonstrate reduced IFNγ ELISpot expression in sepsis patients, especially in those with adverse clinical outcomes (20,21,50). However, in contrast to these previous studies, we used diluted whole blood in our ELISpot assay instead of isolated PBMCs and observed increased IFNγ production in both unstimulated and stimulated whole blood from critically ill patients. There are two key advantages of using diluted whole blood in the ELISpot. First, the use of whole blood permits the assay to take place with the entire blood composition (i.e., all leukocytes, erythrocytes, platelets, and plasma proteins and metabolites) maintained. Responses to critical illness and *ex vivo* stimulation may be either direct or be mediated via cell-cell communication and/or plasma mediators. Traditional processing of blood by density gradient centrifugation separates the PBMC from the neutrophils, platelets, and plasma. Second, it is a simpler and more ‘rapid’ assay to set up because there is no required cell isolation step.

On days 1-7, both the CINS and SEPTIC patients had reduced ALC as compared to healthy controls (**Supplemental Figure 1**). Surprisingly, despite this lymphopenia, the number of lymphocytes spontaneously producing IFNγ was higher in both CINS and SEPSIS patients versus healthy controls at all three time points (**Figure 2**). Day 1 stimulated IFNγ SFU, spot size (SS), and total expression did not differ between cohorts. However, on Day 4, both the number of cells producing IFNγ as well as total expression of IFNγ was higher in both the CINS and SEPSIS cohorts as compared healthy subjects. On Day 7, the number of cells producing IFNγ remained higher in the CINS and SEPSIS cohorts. Of note, the amount of IFNγ produced on a per cell basis (reflected by spot size) was lowest in the SEPSIS cohort with total expression similar to healthy controls.

Given the considerable amount of data showing sepsis can evolve into an immunosuppressed state, it was surprising to see both the spontaneous and stimulated IFNγ production increase in septic patients (early after admission) versus healthy subjects. There are several likely explanations for this apparent paradox. The first potential explanation relates to timing; specifically, the data presented herein came from blood samples collected within the first 7 days after ICU admission. It is difficult to determine exactly when the sepsis-induced hyperinflammation transitions to a state of immunoparalysis, but it is tempting to speculate that our assessment of immune fitness was still within the window of hyperinflammation and exacerbated immune cell activity.

A second explanation has been termed “*bystander activation*” (51,52). The inflammatory response that develops during infection has a capacity to trigger antigen-experienced effector and/or memory CD8^+^ T cells present in a T cell receptor-independent and cytokine-dependent manner. A number of cytokines including IL-12, IL-15, TNFα, and IL-18 induce CD8^+^ T cell activation and resultant IFNγ production (53). Thus, the sepsis cytokine milieu likely primes pre-existing, effector and memory CD8^+^ T cells to produce IFNγ in a cognate antigen-independent fashion (**Figure 2**). In addition, these cytokine-primed effector/memory CD8^+^ T cells will also respond with IFNγ production to a myriad of cytokines produced *ex vivo* during anti-CD3/CD28 stimulation.

A third explanation may lie in the differences in the lymphocyte subsets present in the peripheral blood of healthy subjects versus CINS and SEPSIS patients at the time of blood collection. *De novo* clonal expansion of pathogen-specific effector CD8^+^ T cells in response to sepsis-inducing pathogens and resultant inflammation leads the potential for a preponderance of active response effector cells – especially early in the septic timeline. In contrast, healthy control volunteers are more likely to have ‘resting’ naïve and memory T cells and a minimal (if any) increase in inflammatory cytokines. Consequently, the number of T cells capable of rapidly responding to polyclonal and/or bystander cytokine stimulations and produce IFNγ in the ELISpot assay is increased in SEPSIS patients compared with healthy subjects.

### Current Work

The current studies add to the body of information suggesting ELISpot examining whole blood production of IFNγ can both discriminate long-term mortality and identify those patients who may benefit from therapeutic interventions targeting adaptive immunity.

### Limitations

This study has several limitations. Despite multicenter enrollment, sample sizes were still relatively small for discriminative modeling. Over the past two decades, improved in-hospital management has reduced the number of adverse events and in-hospital mortality to sepsis and critical illness (25,54). Discriminatory analyses could only be conducted in the SEPSIS cohort as CINS patients had very low in-hospital (1%) and 180-day mortality (4%) (**Table 2**). Second, every effort was made to match healthy control subjects to the SEPSIS and CINS cohorts, but the healthy donors used in this study were, as a group, significantly younger and more frequently female (**Table 1**). Median ages in the healthy control cohort were greater than 45 years, a break point often determined to be associated with increased adverse outcomes in critically ill patients (25). Despite the multicenter nature of the study, the cohorts still were also predominantly Caucasian.

### Future Directions

While the findings presented herein suggest assessing IFNγ production by ELISpot can be useful in identifying sepsis patients at risk of long-term mortality and the immune suppressed endotype, its discriminative ability is similar to SOFA and Charlson comorbidity indices and does not add significantly to their discriminative power. With that said, SOFA and Charlson comorbidity indices are rarely used for clinical decision making because they provide no insights or therapeutic directions into the immunological disturbances associated with sepsis and adverse outcomes. Application of ELISpot to the clinical armamentarium has the potential to provide important information regarding which sepsis patients would benefit from targeted therapy (precision medicine). For example, septic patients who have profound suppression of stimulated IFNγ production may be harmed by therapy with corticosteroids, but might be good candidates for immune adjuvants therapies to boost their ability to combat invading pathogens. ELISpot is an FDA-cleared approach for assessing functional immune status to prior tuberculosis infection and the ELISpot reader used in these studies (CTL S6 Entry) is FDA 510(k) cleared. However, to make these results more actionable, ELISpot results will need to be obtained within hours, instead of days. Currently, ELISpot results take at least 24 hours to return, although preliminary data from our consortium suggests the assay can be modified to produce results in less than 12 hours (*Griffith, T and Caldwell, C., manuscript in preparation*).

In addition, ELISpot can be readily used to assess other components of the blood innate and adaptive immune response simply by varying the stimulant and the readout metric. For example, innate immune responses have been readily assessed using endotoxin or other TLR ligands as a whole blood stimulant and TNFα as the readout (20,21). Furthermore, underlying mechanisms of adaptive or innate immune responses can be explored using alternative stimulants (30,31,55), simultaneous adjuvants or inhibitors (56), or different readout metrics (21,55).

## Conclusions

ELISpot can assess functional immune status in critically ill patients, predict adverse long-term outcomes, and identify subsets of patients who may benefit from immunostimulant therapy.

## Contributions

### University of Florida

**EAB** –Investigation, Visualization, Writing-Review and Editing; **PAE**- Resources, Funding Acquisition, Supervision, Writing-Review and Editing; **ML**-Investigation, Visualization; **LEB-** Supervision, Investigation, Writing-Review and Editing; **TL-** Supervision, Investigation, Writing-Review and Editing; **VP**- Investigation, Visualization, **LLM**- Conceptualization, Funding Acquisition, Resources, Supervision, Project Administration, Writing-Review and Editing; **RU**- Supervision, Investigation, Visualization, **JPL**- Supervision, Investigation, **MD**-Investigation, Visualization, **LZS**-Investigation, Visualization, **RD**-Investigation, **AM**-Investigation

### University of Iowa

**EES**– Investigation, Visualization; **MH**– Investigation, Visualization; **PMcG** – Conceptualization, Project Administration; **MR**– Investigation, Visualization; **VPB**– Conceptualization, Funding Acquisition, Resources, Supervision, Writing-Review and Editing

### University of Minnesota

**RWG** – Conceptualization, Project administration; **TSG**– Conceptualization, Funding, Acquisition, Resources, Supervision, Writing–Review and Editing; **LH**– Project Administration; **TAK**– investigation, visualization; **CN**– project administration; **JX**– investigation, visualization.

### Washington University in St. Louis

**RSH-** Conceptualization, Funding, Acquisition, Resources, Supervision, Writing–Review and Editing; **DDT-** Investigation, **AD**- Investigation, **SM1**-Investigation, **SM2**-Investigation, **IT**- Conceptualization, Supervision, Writing-Review and Editing

### University of Washington

**SB**- Conceptualization, Funding, Writing–Review and Editing

### University of Cincinnati

**CCC**- Conceptualization, Funding, Acquisition, Resources, Supervision, Writing–Review and Editing; **MDG**- Acquisition, Investigation, Visualization, Supervision. **CBB** – Acquisition, Investigation

### Case Western Reserve University

**MBM-** Conceptualization, Supervision, Investigation, Writing-Review and Editing; **KR**- Conceptualization, Investigation, Acquisition, Writing-Review and Editing.

## Data Availability

All data produced in the present study are available by request from the following site: https://www.ctsi.ufl.edu/research/laboratory-services/ctsi-biorepository-2/scirc-specimens-archive/

## Abbreviations

ALC: absolute lymphocyte count
AMA: discharged against medical advice
AUROC: area under the receiver-operator curve
CINS: critically ill, nonseptic cohort of patients
CTSI: UF Clinical and Translational Science Institute
ELISpot: enzyme-linked immunospot assay
HLA-DR: human leukocyte antigen DR isotype
IFNγ: interferon-γ
IL-6: interleukin 6
IL-10: interleukin 10
IPR: in-patient rehabilitation
LTAC: long-term acute care facility
mAb: monoclonal antibody
PBMC: peripheral blood mononuclear cell
SEPSIS: critically ill, sepsis cohort of patients
SFU: ELISpot spot forming units
SNF: specialized nursing facility
SOFA: sequential organ failure assessment score
sPD-L1: soluble programmed death ligand-1
SS: ELISpot spot size
TE: ELISpot total expression

## Acknowledgements

This work was directly supported by R01 GM-132364, awarded by the National Institute of General Medical Sciences, including a supplement (R01 GM-132364- 03S1) in 2022-2024.

Additional support was provided by grants T32 GM-008721 (EAB, VP, PAE), RM1 GM-139690 (LLM, PAE), R01 GM-149657 (TL), R01 GM-124156 (MDG), R35 GM-140806 (PAE), R35 GM- 133756 (IT), R35 GM-134880 (VB), R35 GM-134880 (SB), R35 GM1-40881 (TSG), R35 GM-12698 (RSH), all from the National Institute of General Medical Sciences, G102983- 6263608307-1 (MDG) from Department of Defense, IK6BX006192 from the Department of Veteran Affairs (TSG), and BE 7016/1-1 (CD) from the Deutsche Forschungsgemeinschaft (German Research Foundation).

The authors also wish to gratefully acknowledge the patients and their families for their willingness to participate in an observational clinical study with no direct benefit to them.

---

**Supplementary Table 1.**
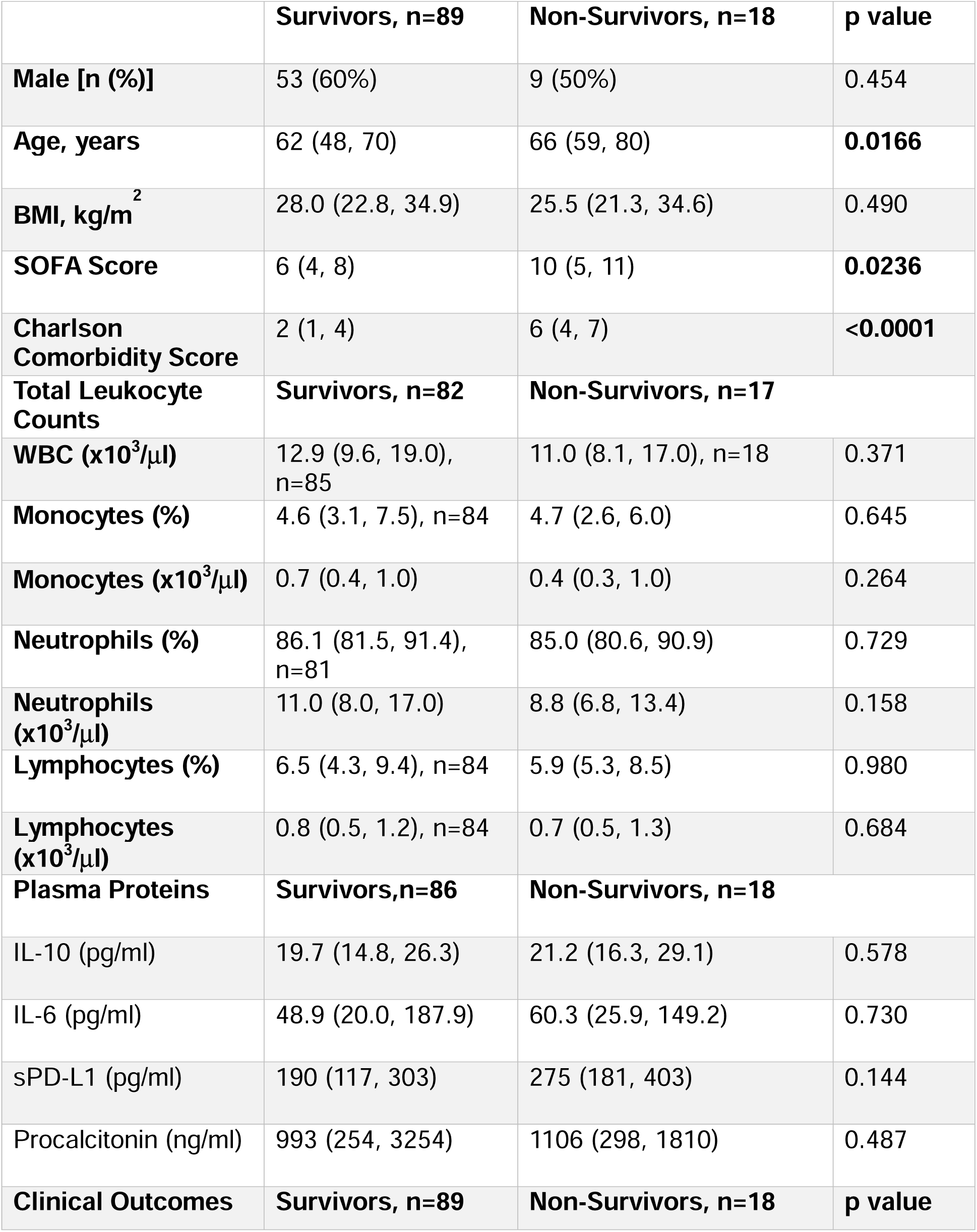
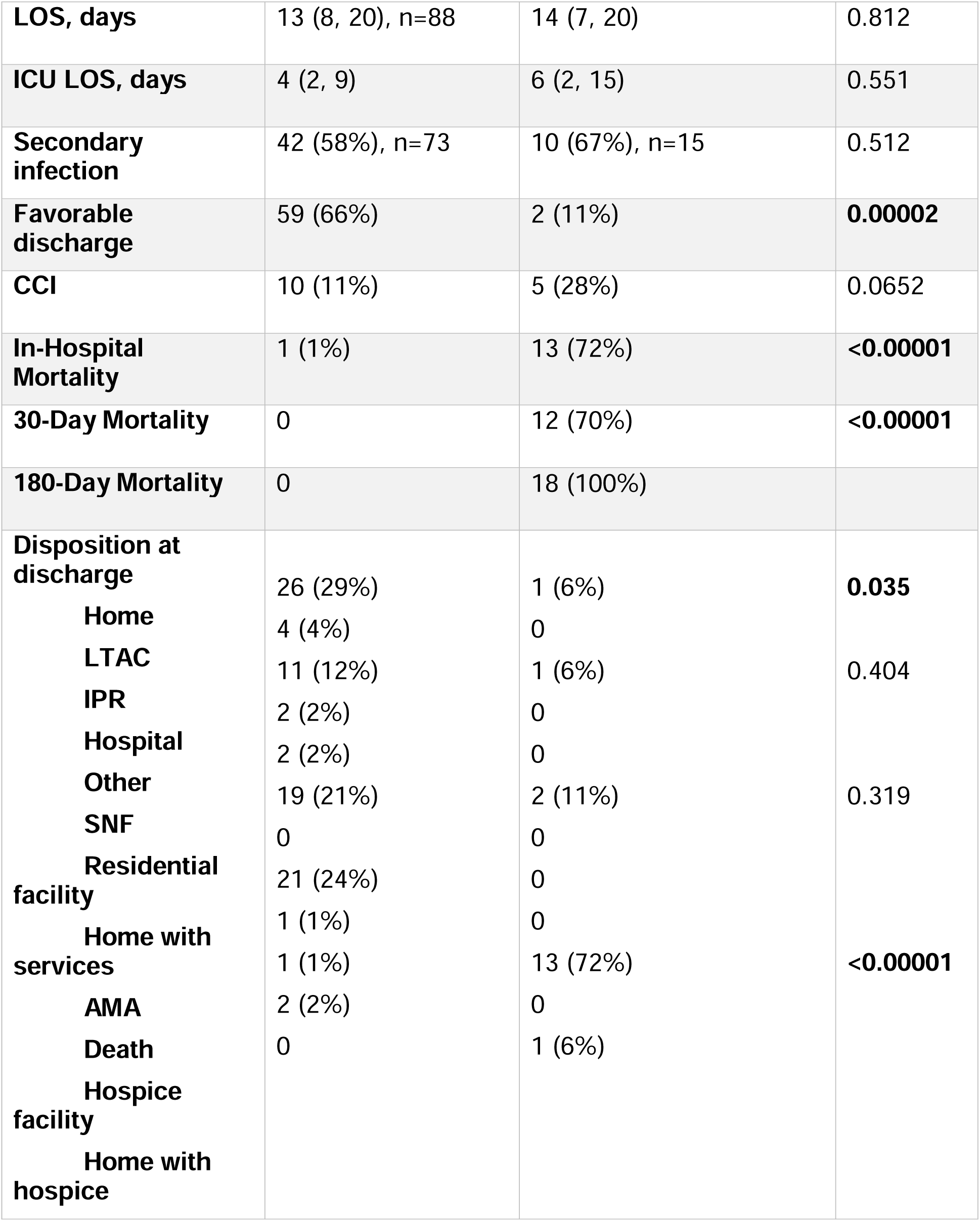
Demographics and Outcomes Between SEPSIS Patients who Survived or Died Within 180 Days after Sepsis. Values represent the number of sample measurements for each analyte.

**Supplemental Figure 1.**
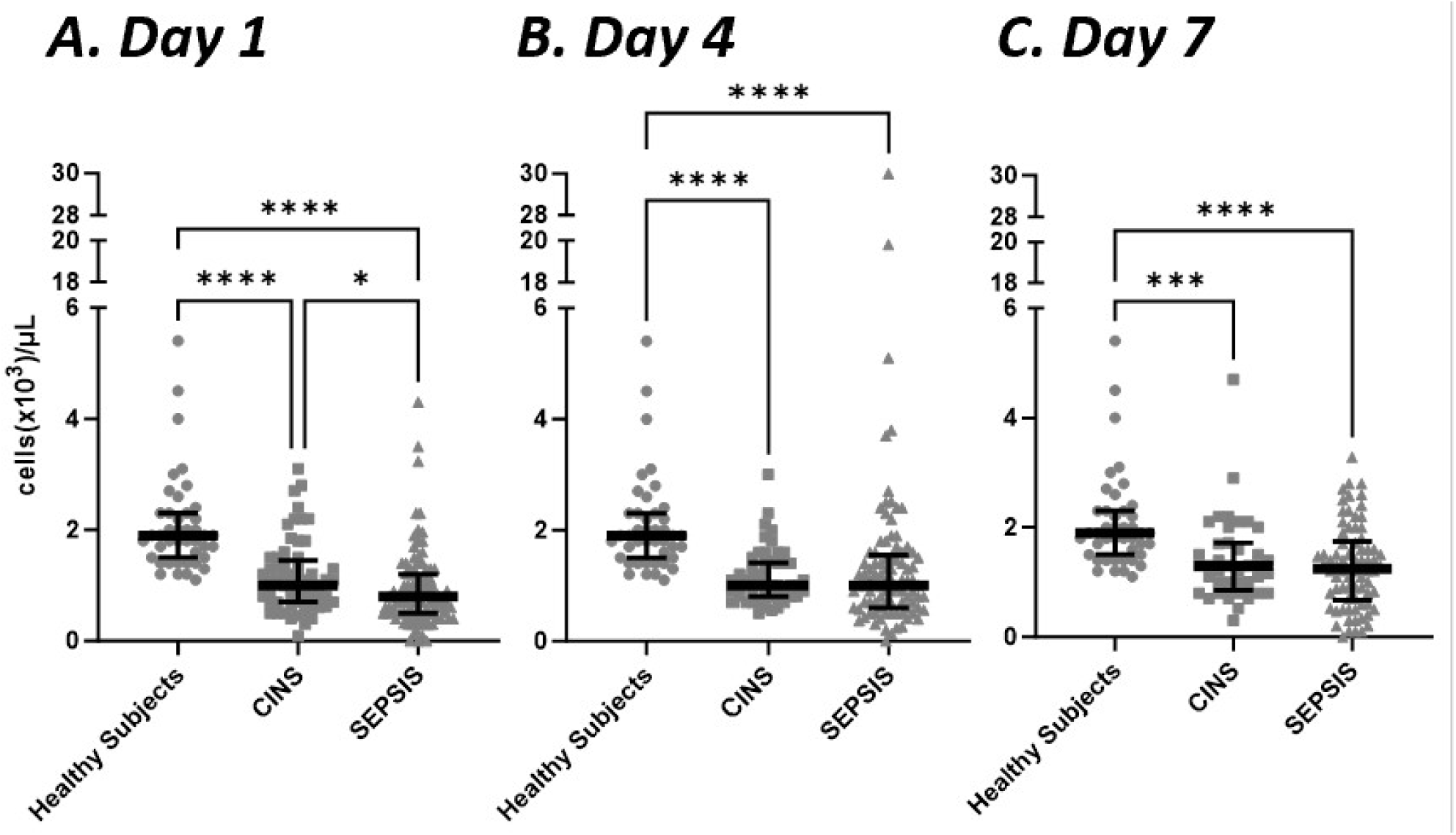
Absolute Lymphocyte Counts in SEPSIS and CINS Cohorts at Different Time Intervals and Healthy Subjects. Whole blood was collected at different time points and total and absolute lymphocyte counts were determined. Both SEPSIS and CINS resulted in a significant decline in total lymphocytes when compared to healthy subjects on days 1, 4 and 7, while absolute lymphocyte counts were lower in SEPSIS than in CINS patients on day 1. Healthy control subjects were sampled only once but values are presented at each time point for comparison. * p<0.05, *** p<0.001, **** p<0.0001 as determined by Kruskal-Wallace ANOVA and post-hoc analyses using the Dunn test. Values are two sided and represent raw p values.

**Supplemental Figure 2.**
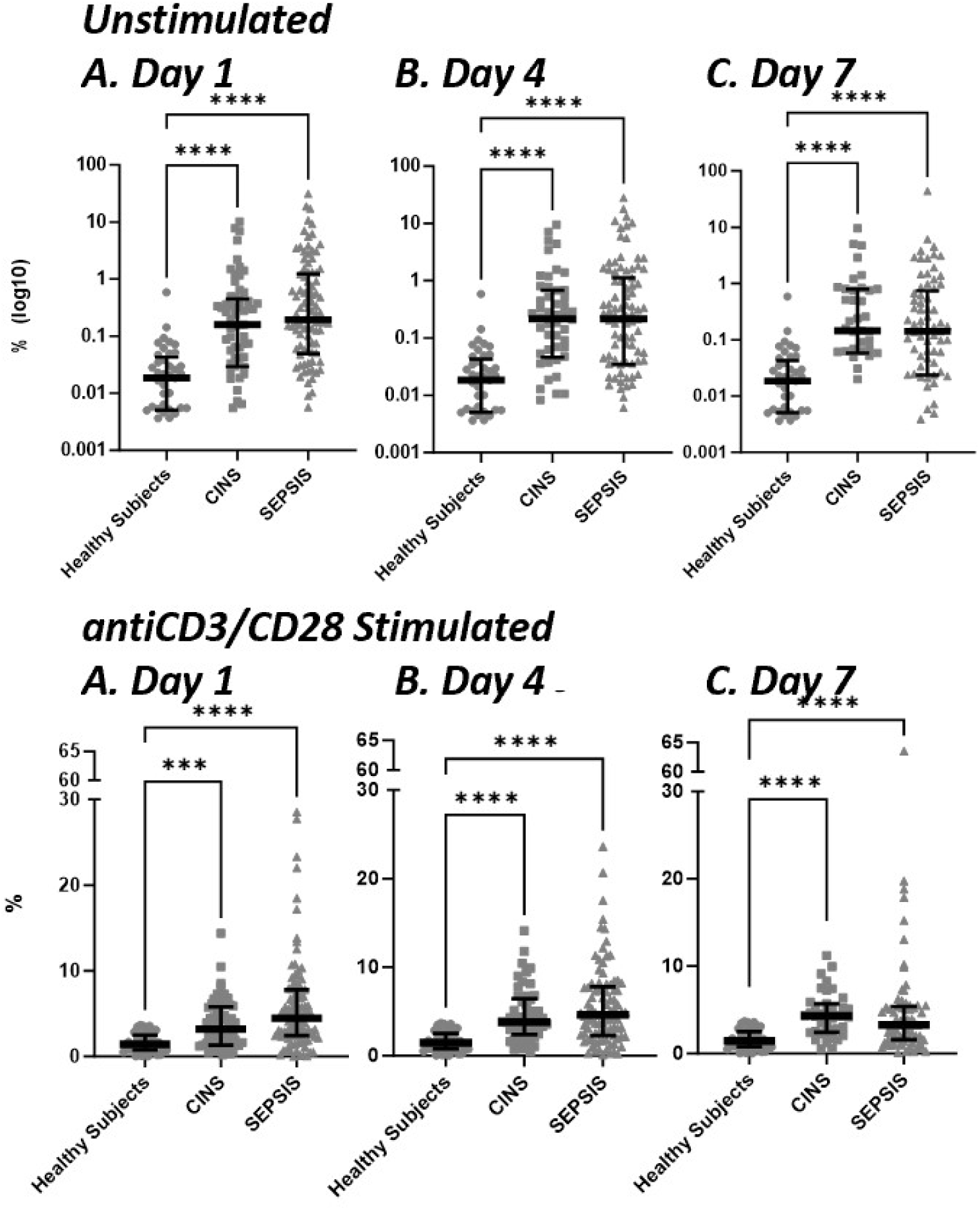
Percentage of Lymphocytes Expressing IFNγ in Whole Blood from SEPSIS and CINS cohorts, and Healthy Subjects. The number of spot forming units was compared to the absolute lymphocyte count and the percentage of lymphocytes producing IFNγ was calculated. Regardless of time after enrollment, both SEPSIS and CINS markedly increased the percentage of IFNγ-producing cells. * p<0.05, *** p<0.001, **** p<0.0001, as determined by Kruskal-Wallace ANOVA and post-hoc analyses using the Dunn test. Values are two sided and represent raw p values. SFU, spot forming units. SS, spot size. TE, total IFNγ expression.

## Supplemental Methods

Detailed specific criteria for consenting and enrolling SEPSIS and CINS patients, and healthy control subjects are described below, as is sampling and processing procedures. Actual IRB documentation and the Consortium Laboratory Manual are readily available by simply requesting them from our data and sample broker, the UF Clinical and Translational Research Institute Biorepository (https://www.ctsi.ufl.edu/research/laboratory-services/ctsi-biorepository-2/scirc-specimens-archive/).

### Recruitment methods

Screening for sepsis will be carried out using each hospital’s own version of their sepsis alert system, which quantifies derangements in vital signs, white blood cell count, and mental status. After a putative diagnosis of sepsis, the patient is transferred to the ICU and sepsis treatment bundles are initiated. If a patient is believed to have an infection and they are located in, or transferred to the ICU, they are entered into each institution’s sepsis management protocol as standard of care which implements a variety of standard operating procedures (SOPs) of clinical ICU care. After the patient enters the clinical management protocol, the research coordinator is notified of a potential research subject. This coordinator will then assess the following inclusion/exclusion criteria and consent the appropriate candidates.

All critically ill patients with or without sepsis will be managed via each institution’s evidence-based management protocols that emphasize early antibiotic administration, fluid resuscitation and hemodynamic monitoring and support, consistent with current Surviving Sepsis Campaign guidelines.

### Consenting

Consent will be sought by clinical research staff, all of whom are familiar with institutional logistics and infrastructure, sample acquisition and preparation, and are experienced in the nuances of enrollment and informed consent for this challenging patient population. Many critically ill patients in the ICUs may have altered mental status or pharmacologic sedation, but would be regarded as prospective research subjects. We will seek an IRB-approved 96-hour delayed consent for blood sample acquisition and completion of the /T1 visit per protocol (within the first 72 hours). This includes: study criteria evaluation, sample collection and transportation to clinical and research laboratories, collection of demographic information, and collection of medical laboratory results to compliment research experiments. Additionally, we will seek approval for telephone consent in the event that no LAR is physically present. For the ICU patient population, many of whom are pharmacologically sedated and mechanically ventilated, initial consent is commonly requested from LAR/next of kin. The setting for conveying consent information to LAR/next of kin is often the ICU family waiting area or at bedside. Consideration is given to the emotional status of the LAR/next of kin and ability to understand the basic intent, methods, and voluntary nature of participation. If we need to call a LAR, ideally we will email them a copy of the consent and review, record approval with a witness on the line. If we can’t email them, then we will review the entire consent over the phone, record approval with a witness on the line.

The research staff will monitor the patient’s progress and once they regain capacity, the patient will be re-consented. If patient does not regain capacity to consent by discharge, he/she will be consented at their next follow-up visit if capacity is regained.

For the healthy control subjects, we will send an email out to the Department of Surgery email distribution list to all department staff for recruiting healthy control participants. When the participant comes in for the blood draw we will review the consent and have them sign.

### Specific Inclusion/Exclusion Criteria

### SEPSIS-Presumed Sepsis Patients

#### Inclusion Criteria

A. Directly admitted ICU patients with sepsis (From ED or OR) ICU patient developing sepsis during hospitalization. Transferred to ICU from inpatient unit for development of sepsis. Documentation in notes or diagnoses of "Sepsis", "Septic shock", "Severe Sepsis"

#### or

B. Suspicion of infectious cause of illness on admission Documentation of suspected infection Infectious testing performed (blood, urine, other cultures, viral or fungal testing) Diagnostic testing: chest X-ray. CT scan of abdomen Initiation of antimicrobial therapy (antibiotics, antivirals, antifungals) Source control operation/procedure performed

#### and

Organ dysfunction as defined as an acute change in total SOFA score of 2 points consequent to the infection.
The baseline SOFA score can be assumed to be zero in patients not known to have preexisting organ dysfunction.

#### Exclusion Criteria

Patients deemed to be futile care or have advanced care directives or goals of care limiting resuscitative efforts.
Severe traumatic brain injury (evidence of neurologic injury on CT scan and a GCS <8 after resuscitation).
Refractory shock (i.e., patients who are expected to die within 24hours).
Uncontrollable source of sepsis (e.g. irreversible disease state such as unresectable dead bowel).
Outside facility transfer where suspected sepsis onset is >72 hours prior to ICU admission.
Active chronic hepatitis or other chronic infectious diseases.
Known HIV infection with CD4 count <200 cells/mm^3^.
Organ transplant recipient on immunosuppressive agents.
Known pregnancy.
Prisoners.
Institutionalized patients.
Active cancer.
Any recent (past 6 months) chemotherapy or immunomodulatory therapies (including biologics, monoclonal antibodies).
Patient having received steroids in past 4 weeks.

### CINS – Critically-ill non-sepsis patients

#### Inclusion Criteria

A. Patients admitted to the SICU for non-infectious reasons Trauma patients Post-operative patients (not infectious source control procedures) Inpatients transferred to the SICU for non-infectious reasons (e.g., bleeding, volume overload, acute cardiac issue, ect.)

#### Exclusion criteria

Patients deemed to be futile care or have advanced care directives or goals of care limiting resuscitative efforts.

Severe traumatic brain injury (evidence of neurologic injury on CT scan and a GCS <8 after resuscitation).

Refractory shock (i.e., patients who are expected to die within 24hours).

Uncontrollable source of sepsis (e.g. irreversible disease state such as unresectable dead bowel).

Outside facility transfer where suspected sepsis onset is >72 hours prior to ICU admission.

Active chronic Hepatitis or other chronic infectious diseases.

Known HIV infection with CD4 count <200 cells/mm3.

Organ transplant recipient on immunosuppressive agents.

Known pregnancy.

Prisoners.

Institutionalized patients.

Active cancer.

Any recent (past 6 months) chemotherapy or immunomodulatory therapies (including biologics, monoclonal antibodies).

Patient having received steroids in past 4 weeks.

### Healthy Control Subjects

#### Inclusion Criteria

All adults (age >=18)

Ability to obtain Informed Consent prior to blood collection.

#### Exclusion Criteria

Current, chronic steroid use

Pregnancy

Current or recent (within 7 days) use of antibiotics.

### Sample Collection

**The overview of research sample collection for SPIES clinical study are presented in the Sample Collection Chart:**

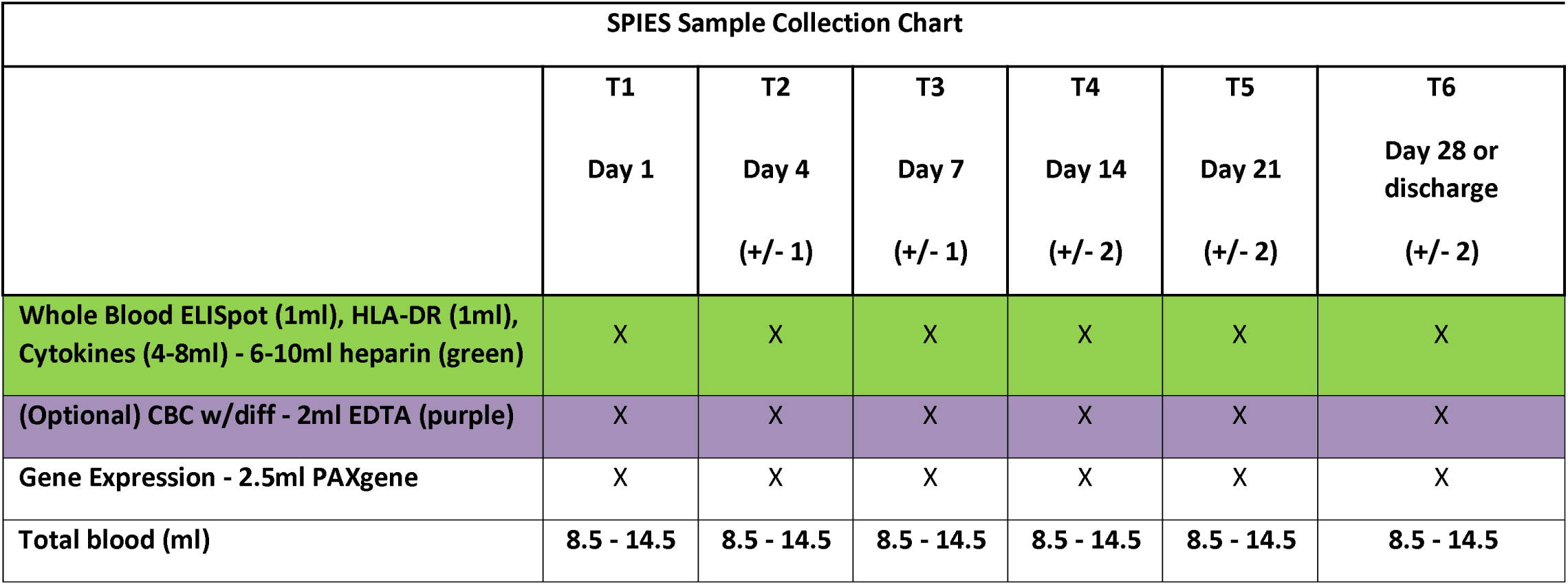

| SPIES Sample Collection Chart |  |  |  |  |  |  |
| --- | --- | --- | --- | --- | --- | --- |
|  | T1<br>Day 1 | T2<br>Day 4<br>(+/- 1) | T3<br>Day 7<br>(+/- 1) | T4<br>Day 14<br>(+/- 2) | T5<br>Day 21<br>(+/- 2) | T6<br>Day 28 or<br>discharge<br>(+/- 2) |
| Whole Blood ELISpot (1ml), HLA-DR (1ml),<br>Cytokines (4-8ml) - 6-10ml heparin (green) | X | X | X | X | X | X |
| (Optional) CBC w/diff - 2ml EDTA (purple) | X | X | X | X | X | X |
| Gene Expression - 2.5ml PAXgene | X | X | X | X | X | X |
| Total blood (ml) | 8.5 - 14.5 | 8.5 - 14.5 | 8.5 - 14.5 | 8.5 - 14.5 | 8.5 - 14.5 | 8.5 - 14.5 |

### PLASMA PROCESSING

Administrative Note: All personnel involved in the procedures are to have completed the University required Blood-Borne Pathogen training program, and provided suitable personal protection equipment (PPE). These procedures are to be performed whenever possible in a biocontainment hood (BSL1 or BSL2). All solid or liquid waste derived from the blood must be disposed of appropriately in a biohazard container or inactivated using bleach or an acceptable disinfectant. Venous or arterial whole blood is collected into the appropriate blood collection tubes for each time point. The collection of blood should be obtained from an existing arterial or venous line, or venipuncture should be performed by someone experienced in the technique, and familiar with infectious precautions. The blood should be processed as soon as possible, **<u>but within 3 hours of the draw</u>**. The blood should be **<u>kept cold on ice</u>** during the period from the draw to the initiation of processing.

### CBC with Differential

Each site must collect for and obtain a CBC with Differential at every patient sample time point. These must be collected at the same time as the study blood draws or as close to it as possible if done through the hospital’s standard of care. This data must be entered into REDCap.

### Plasma Collection

1. Invert the green heparin blood tube gently to thoroughly mix the blood. Transfer one ml of blood to a 15 ml conical (polyethylene or polystyrene) tube labeled HLA-DR. And transfer 500 ul of blood to a 1.5 ml microcentrifuge tube labeled ELISpot.
2. After removing the blood needed for the ELISpot and Smart Tube assays in Step 1, centrifuge the remaining blood in the green top heparin blood collection tube at room temperature (22°C) for 10 minutes at 1,800 x g, with the brake on low.
3. Once the tube has finished spinning, carefully transfer the plasma from the green top blood tube (take care not to disturb the cellular constituents) and aliquot 500 ul of plasma equally into 4-6 **green** capped plasma collection tubes. If there is any remaining plasma, collect and distribute evenly among all tubes.
4. Make sure the appropriate *Heparin Plasma* barcode labels are attached to the tubes and immediately store at -80°C.

## Notes

**Conflict of Interest Statements:** Evan A, Barrios, M.D. reports institutional support for salary from a National Institute of General Medical Sciences training grant in burns, trauma and sepsis (T32 GM-008721) (PAE). Monty B. Mazer, M.D. is a member of Immune Functional Diagnostics, LLC and receives no direct financial compensation. Immune Functional Diagnostics, LLC is developing predictive metrics in critical illness and this technology is evaluated in this research. Patrick McGonagill M.D. reports no conflicts of interest. Christian B. Bergmann, M.D. is supported by grants from the Deutsche Forschungsgemeinschaft (German Research Foundation) (BE 7016/1-1). Michael D. Goodman, M.D. reports institutional support for salary from the National Institute of General Medical Sciences (R01 GM-124156) and the Department of Defense (G102983-6263608307-1). Robert W. Gould, M.D. reports no conflicts of interest. Mahil Rao M.D., Ph.D. reports no conflicts of interest. Valerie Polcz, M.D. reports institutional support for salary from a National Institute of General Medical Sciences training grant in burns, trauma and sepsis (T32 GM-008721) (PAE). Ruth Davis, B.S.N, R.N. reports no conflicts of interest. Drew Del Toro, M.D. reports no conflicts of interest. Marvin Dirain, M.S. reports no conflicts of interest. Alexandra Dram reports no conflicts of interest. Lucas Hale, B.A. reports no conflicts of interest. Mohammad Heidarian, M.S. reports no conflicts of interest. Tamara A. Kucaba, B.S. reports no conflicts of interest. Jennifer P. Lanz, M.S.N., R.N. reports no conflicts of interest. Ashley McCray, R.N. reports no conflicts of interest. Sandra Meszaros reports no conflicts of interest. Sydney Miles, B.S. reports no conflicts of interest. Candace Nelson, B.A. reports no conflicts of interest. Ivanna Rocha, M.P.H. reports no conflicts of interest. Elvia E Silva, M.S. reports no conflicts of interest. Ricardo Ungaro, B.S. reports no conflicts of interest. Andrew Walton, M.S. reports no conflicts of interest. Julie Xu, B.S. reports no conflicts of interest. Leilani Zeumer-Spataro, B.S. reports no conflicts of interest. Muxuan Liang, Ph.D. reports no conflicts of interest. Isaiah Turnbull, M.D is a member of Immune Functional Diagnostics, LLC and receives no direct financial compensation. Immune Functional Diagnostics, LLC is developing predictive metrics in critical illness and this technology is evaluated in this research. He is also supported by R35 GM-133756 from the National Institute of General Medical Sciences. Letitia E. Bible, M.D. reports no conflict of interest. Tyler Loftus, M.D. is supported by R01 GM-149657 from the National Institute of General Medical Sciences. Philip A. Efron, M.D. is supported by grants R35 GM-140806, T32 GM-008721 and RM GM- 139690 awarded by the National Institute of General Medical Sciences. Kenneth E. Remy, M.D., M.S. is a member of Immune Functional Diagnostics, LLC and receives no direct financial compensation. Immune Functional Diagnostics, LLC is developing predictive metrics in critical illness and this technology is evaluated in this research. Scott Brakenridge, M.D., M.S.C.S. is funded by R35 GM-134880 from the National Institute of General Medical Sciences. Dr. Brakenridge and the University of Florida may receive royalty income based on a technology developed by Dr. Brakenridge and others and licensed by Washington University in St. Louis to IFDx LLC. That technology is evaluated in this research. Vladimir P. Badovinac Ph.D.is funded by R35 GM-134880 from the National Institute of General Medical Sciences. Thomas S. Griffith, Ph.D. is funded by R35 GM-140881 from the National Institute of General Medical Sciences and the recipient of a Research Career Scientist award (IK6BX006192) from the Department of Veteran Affairs. Lyle L. Moldawer, Ph.D. is supported by NIH grants RM1 GM-139690, R01GM-132364 and RF1 NS128626. Dr. Moldawer and the University of Florida may receive royalty income based on a technology developed by Dr. Moldawer and others and licensed by Washington University in St. Louis to IFDx LLC. That technology is evaluated in this research. Richard S. Hotchkiss, M.D. Dr. Hotchkiss and Washington University in St. Louis may receive royalty income based on a technology developed by Dr. Hotchkiss and others and licensed by Washington University in St. Louis to IFDx LLC. That technology is evaluated in this research. He is also supported by R35 GM-126928, awarded by the National Institute of General Medical Sciences. Charles C. Caldwell, Ph.D. Dr. Caldwell and the University of Cincinnati may receive royalty income based on a technology developed by Dr. Caldwell and others and licensed by Washington University in St. Louis to IFDx LLC. That technology is evaluated in this research.

### Competing Interest Statement

Evan A, Barrios, M.D. reports institutional support for salary from a National Institute of General Medical Sciences training grant in burns, trauma and sepsis (T32 GM-008721) (PAE).
Monty B. Mazer, M.D. is a member of Immune Functional Diagnostics, LLC and receives no direct financial compensation. Immune Functional Diagnostics, LLC is developing predictive metrics in critical illness and this technology is evaluated in this research.
Christian B. Bergmann, M.D. is supported by grants from the Deutsche Forschungsgemeinschaft (German Research Foundation) (BE 7016/1-1).
Michael D. Goodman, M.D. reports institutional support for salary from the National Institute of General Medical Sciences (R01 GM-124156) and the Department of Defense (G102983-6263608307-1).
Valerie Polcz, M.D. reports institutional support for salary from a National Institute of General Medical Sciences training grant in burns, trauma and sepsis (T32 GM-008721) (PAE).
Isaiah Turnbull, M.D is a member of Immune Functional Diagnostics, LLC and receives no direct financial compensation. Immune Functional Diagnostics, LLC is developing predictive metrics in critical illness and this technology is evaluated in this research. He is also supported by R35 GM-133756 from the National Institute of General Medical Sciences.
Tyler Loftus, M.D. is supported by R01 GM-149657 from the National Institute of General Medical Sciences.
Philip A. Efron, M.D. is supported by grants R35 GM-140806, T32 GM-008721 and RM GM-139690 awarded by the National Institute of General Medical Sciences.
Kenneth E. Remy, M.D., M.S. is a member of Immune Functional Diagnostics, LLC and receives no direct financial compensation. Immune Functional Diagnostics, LLC is developing predictive metrics in critical illness and this technology is evaluated in this research.
Scott Brakenridge, M.D., M.S.C.S. is funded by R35 GM-134880 from the National Institute of General Medical Sciences. Dr. Brakenridge and the University of Florida may receive royalty income based on a technology developed by Dr. Brakenridge and others and licensed by Washington University in St. Louis to IFDx LLC. That technology is evaluated in this research.
Vladimir P. Badovinac Ph.D.is funded by R35 GM-134880 from the National Institute of General Medical Sciences.
Thomas S. Griffith, Ph.D. is funded by R35 GM-140881 from the National Institute of General Medical Sciences and the recipient of a Research Career Scientist award (IK6BX006192) from the Department of Veteran Affairs.
Lyle L. Moldawer, Ph.D. is supported by NIH grants RM1 GM-139690, R01GM-132364 and RF1 NS128626. Dr. Moldawer and the University of Florida may receive royalty income based on a technology developed by Dr. Moldawer and others and licensed by Washington University in St. Louis to IFDx LLC. That technology is evaluated in this research.
Richard S. Hotchkiss, M.D. Dr. Hotchkiss and Washington University in St. Louis may receive royalty income based on a technology developed by Dr. Hotchkiss and others and licensed by Washington University in St. Louis to IFDx LLC. That technology is evaluated in this research. He is also supported by R35 GM-126928, awarded by the National Institute of General Medical Sciences.
Charles C. Caldwell, Ph.D. Dr. Caldwell and the University of Cincinnati may receive royalty income based on a technology developed by Dr. Caldwell and others and licensed by Washington University in St. Louis to IFDx LLC. That technology is evaluated in this research.
No other relationships or activities that could appear to have influenced the submitted work.

### Clinical Protocols

https://www.ctsi.ufl.edu/research/laboratory-services/ctsi-biorepository-2/scirc-specimens-archive/

### Author Declarations

The Institutional review board (IRB-01) at the University of Florida gave ethical approval for this work (IRB# 202000924, approval 6/5/2020). Under NIH guidelines, the University of Florida IRB served as the sponsoring IRB for this study, and IRBs at the University of Cincinnati and Washington University of St. Louis ceded oversight of the study to the University of Florida.

### Summary of Updates

Additional Author was added (Letitia E. Bible). Number of SEPSIS patients who expired within 180 days was corrected from 20 to 18. Text and tables have been corrected.

