## Supplemental Figures, Tables and Methods for "Adverse Long-Term Outcomes and an Immune Suppressed Endotype in Sepsis Patients with Reduced Interferon-γ ELISpot: A Multicenter, Prospective Observational Study"

| **SPIES Sample Collection Chart** | | | | | | |
| --- | --- | --- | --- | --- | --- | --- |
|  | **T1** | **T2** | **T3** | **T4** | **T5** | **T6** |
|  | **Day 1** | **Day 4** | **Day 7** | **Day 14** | **Day 21** | **Day 28 or discharge** |
|  |  | **(+/- 1)** | **(+/- 1)** | **(+/- 2)** | **(+/- 2)** | **(+/- 2)** |
| **Whole Blood ELISpot (1ml), HLA-DR (1ml), Cytokines (4-8ml) - 6-10ml heparin (green)** | X | X | X | X | X | X |
| **(Optional) CBC w/diff - 2ml EDTA (purple)** | X | X | X | X | X | X |
| **Gene Expression - 2.5ml PAXgene** | X | X | X | X | X | X |
| **Total blood (ml)** | **8.5 - 14.5** | **8.5 - 14.5** | **8.5 - 14.5** | **8.5 - 14.5** | **8.5 - 14.5** | **8.5 - 14.5** |

**Supplementary Table 1. Demographics and Outcomes Between SEPSIS Patients who Survived or Died Within 180 Days after Sepsis.** Values represent the number of sample measurements for each analyte.

|  | Survivors, n=89 | Non-Survivors, n=18 | p value |
| --- | --- | --- | --- |
| **Male [n (%)]** | 53 (60%) | 9 (50%) | 0.454 |
| **Age, years** | 62 (48, 70) | 66 (59, 80) | **0.0166** |
| **BMI, kg/m^2^** | 28.0 (22.8, 34.9) | 25.5 (21.3, 34.6) | 0.490 |
| **SOFA Score** | 6 (4, 8) | 10 (5, 11) | **0.0236** |
| **Charlson Comorbidity Score** | 2 (1, 4) | 6 (4, 7) | **<0.0001** |
| **Total Leukocyte Counts** | **Survivors, n=82** | **Non-Survivors, n=17** | |
| **WBC (x10^3^/μl)** | 12.9 (9.6, 19.0), n=85 | 11.0 (8.1, 17.0), n=18 | 0.371 |
| **Monocytes (%)** | 4.6 (3.1, 7.5), n=84 | 4.7 (2.6, 6.0) | 0.645 |
| **Monocytes (x10^3^/μl)** | 0.7 (0.4, 1.0) | 0.4 (0.3, 1.0) | 0.264 |
| **Neutrophils (%)** | 86.1 (81.5, 91.4), n=81 | 85.0 (80.6, 90.9) | 0.729 |
| **Neutrophils (x10^3^/μl)** | 11.0 (8.0, 17.0) | 8.8 (6.8, 13.4) | 0.158 |
| **Lymphocytes (%)** | 6.5 (4.3, 9.4), n=84 | 5.9 (5.3, 8.5) | 0.980 |
| **Lymphocytes (x10^3^/μl)** | 0.8 (0.5, 1.2), n=84 | 0.7 (0.5, 1.3) | 0.684 |
| **Plasma Proteins** | **Survivors,n=86** | **Non-Survivors, n=18** | |
| IL-10 (pg/ml) | 19.7 (14.8, 26.3) | 21.2 (16.3, 29.1) | 0.578 |
| IL-6 (pg/ml) | 48.9 (20.0, 187.9) | 60.3 (25.9, 149.2) | 0.730 |
| sPD-L1 (pg/ml) | 190 (117, 303) | 275 (181, 403) | 0.144 |
| Procalcitonin (ng/ml) | 993 (254, 3254) | 1106 (298, 1810) | 0.487 |
| **Clinical Outcomes** | **Survivors, n=89** | **Non-Survivors, n=18** | **p value** |
| **LOS, days** | 13 (8, 20), n=88 | 14 (7, 20) | 0.812 |
| **ICU LOS, days** | 4 (2, 9) | 6 (2, 15) | 0.551 |
| **Secondary infection** | 42 (58%), n=73 | 10 (67%), n=15 | 0.512 |
| **Favorable discharge** | 59 (66%) | 2 (11%) | **0.00002** |
| **CCI** | 10 (11%) | 5 (28%) | 0.0652 |
| **In-Hospital Mortality** | 1 (1%) | 13 (72%) | **<0.00001** |
| **30-Day Mortality** | 0 | 12 (70%) | **<0.00001** |
| **180-Day Mortality** | 0 | 18 (100%) |  |
| **Disposition at discharge**  **Home**  **LTAC**  **IPR**  **Hospital**  **Other**  **SNF**  **Residential facility**  **Home with services**  **AMA**  **Death**  **Hospice facility**  **Home with hospice** | 26 (29%)  4 (4%)  11 (12%)  2 (2%)  2 (2%)  19 (21%)  0  21 (24%)  1 (1%)  1 (1%)  2 (2%)  0 | 1 (6%)  0  1 (6%)  0  0  2 (11%)  0  0  0  13 (72%)  0  1 (6%) | **0.035**  0.404  0.319  **<0.00001** |

**Supplemental Figure 1. Absolute Lymphocyte Counts in SEPSIS and CINS Cohorts at Different Time Intervals and Healthy Subjects.** Whole blood was collected at different time points and total and absolute lymphocyte counts were determined. Both SEPSIS and CINS resulted in a significant decline in total lymphocytes when compared to healthy subjects on days 1, 4 and 7, while absolute lymphocyte counts were lower in SEPSIS than in CINS patients on day 1. Healthy control subjects were sampled only once but values are presented at each time point for comparison. * p<0.05, *** p<0.001, **** p<0.0001 as determined by Kruskal-Wallace ANOVA and post-hoc analyses using the Dunn test. Values are two sided and represent raw p values.

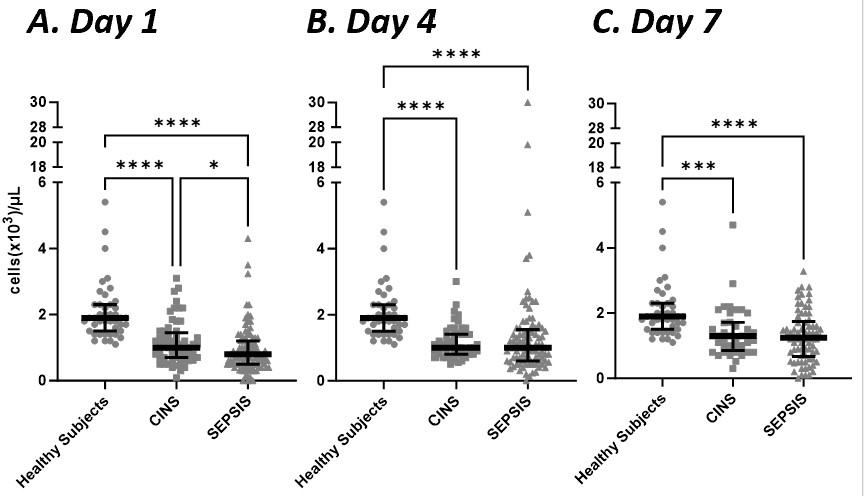

**Supplemental Figure 2. Percentage of Lymphocytes Expressing IFNγ in Whole Blood from SEPSIS and CINS cohorts, and Healthy Subjects**. The number of spot forming units was compared to the absolute lymphocyte count and the percentage of lymphocytes producing IFNγ was calculated. Regardless of time after enrollment, both SEPSIS and CINS markedly increased the percentage of IFNγ-producing cells. * p<0.05, *** p<0.001, **** p<0.0001, as determined by Kruskal-Wallace ANOVA and post-hoc analyses using the Dunn test. Values are two sided and represent raw p values. SFU, spot forming units. SS, spot size. TE, total IFNγ expression.

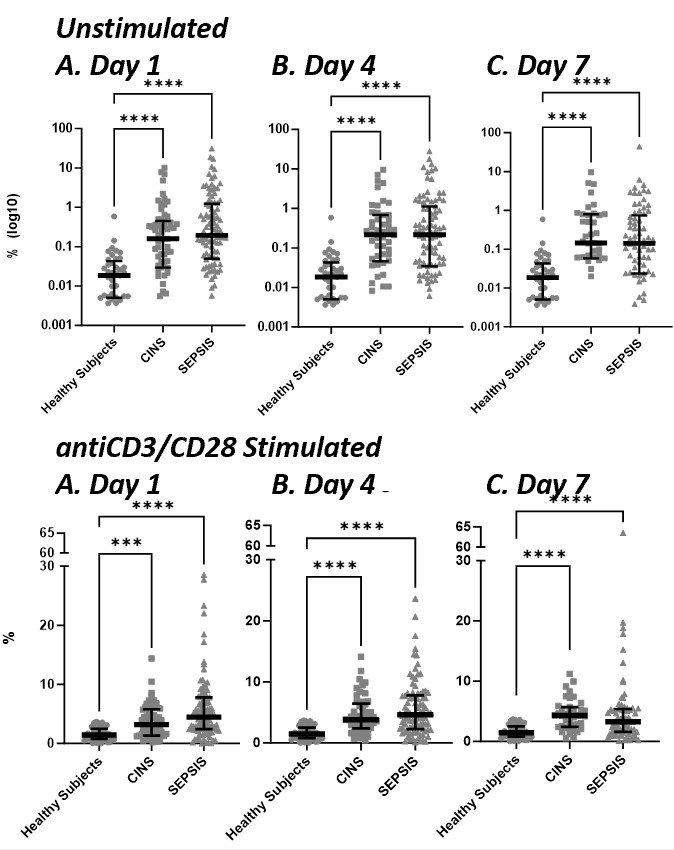
